# Lithium therapy and delayed progression of Alzheimer’s disease and related dementias in patients with bipolar disorder and mild neurocognitive disorders

**DOI:** 10.64898/2026.01.12.26343472

**Authors:** Andrew R. Weckstein, Sinclair Carr, Philip Wang, Nils Krüger, Goodarz Danaei, Sebastian Schneeweiss, Rishi J. Desai

## Abstract

Recent mechanistic evidence identifies lithium deficiency as a driver of Alzheimer’s disease and related dementias (ADRD) pathogenesis. To evaluate this hypothesis in a population-based setting, we emulated a target trial comparing ADRD progression after initiation of lithium versus antiepileptic mood stabilizers among adults ≥55 years with bipolar disorder and mild neurocognitive disorder, using US Medicare claims with replication in two commercial databases.

Lithium initiation was associated with a lower 5-year risk of progression to advanced ADRD (risk ratio 0.87; 95% CI 0.78-0.99) and long-term care stay with ADRD (0.75; 0.56-0.97). Lower risks were most pronounced in patients with later stages of mild cognitive impairment. Results were consistent across sensitivity analyses and replications. Validations using linked cognitive assessments from standardized instruments strengthened confidence in findings. While results may be susceptible to residual bias, this study supports investigating lithium’s potential to delay ADRD progression with randomized trials of optimized lithium formulations.

## Main text

Recent mechanistic evidence suggests endogenous lithium plays a key role in the pathogenesis of Alzheimer’s disease and related dementias (ADRD).^1^ In mouse models, reducing brain lithium accelerated amyloid-β deposition, tau hyperphosphorylation, neuroinflammation, and neuronal loss, causing cognitive decline. Lithium orotate treatment prevented and reversed these changes in mice and restored cognition, supporting lithium supplementation as a potential strategy for addressing ADRD-related cognitive decline.

However, clinical evidence of lithium’s effects on ADRD progression remains limited and contradictory. Initial meta-analyses of three small randomized trials found lithium supplementation to be associated with reduced cognitive impairment in persons with Alzheimer’s disease,^2,3^ but a more recent meta-analysis reported no difference in cognitive decline for lithium relative to placebo.^4^ Population-based observational studies have yielded similarly mixed results.^5–9^ Prior population-based studies have exclusively assessed new-onset dementia incidence rather than disease progression, which makes their findings clinically less compelling regarding lithium’s disease-modifying potential. Further, most studies used designs susceptible to bias from suboptimal comparator selection and other methodological limitations.

Lithium, administered clinically as carbonate or citrate, is a long-standing first-line maintenance treatment for bipolar disorder.^10,11^ Routine clinical use of lithium among patients with co-existing bipolar disorder and mild neurocognitive disorders (MND) presents a unique opportunity to evaluate whether lithium therapy slows cognitive decline in real-world populations.

To that end, we emulated a new-user, active comparator target trial^12^ to compare ADRD progression after initiation of lithium versus antiepileptic mood stabilizers in patients with bipolar disorder and MND.^13^ ADRD follows a progressive clinical continuum from prodromal symptoms through mild cognitive impairment (MCI) to established dementia of increasing severity.^14,15^ We defined MND eligibility criteria to encompass the early-to-middle stages of this continuum, including age-related cognitive decline or memory loss (representing suspected prodromal disease), MCI, or mild-moderate ADRD without complications. Main outcomes included progression to advanced ADRD, operationalized as an incident diagnosis for severe or complicated ADRD, and a long-term care stay (at least 100 days) with an ADRD diagnosis. We used US Medicare claims (2013-2022) for a random 20% sample of enrollees and replicated analyses by emulating the same target trial (**Extended Data Table 1**) in Merative MarketScan (2013-2023) and Optum Clinformatics Data Mart (CDM; 2004-2025) claims databases. The study design and analyses were pre-registered^16^ (**Table S2**).

## Results

A total of 9,461 Medicare beneficiaries met eligibility criteria, including 1,133 lithium initiators and 8,328 antiepileptic mood stabilizer initiators (**Fig. S2**). The average duration of enrollment in Medicare before baseline was 4.9 years in both treatment groups. Time from first MND diagnosis to treatment initiation averaged 15 months for lithium initiators and 16 months for antiepileptic mood stabilizer initiators.

Before adjustment for potential baseline confounders, both treatment groups had a mean age of 69 years at initiation. Psychiatric hospitalizations and other markers of severe bipolar disorder were more common in patients who initiated lithium compared to patients who initiated antiepileptic mood stabilizers (**Table 1; Table S6**). In contrast, lithium initiators had fewer comorbidities and less frailty, which may partly reflect clinical caution in prescribing lithium in patients with severe kidney or cardiovascular disease. Lithium initiators had slightly lower prevalence of mild-moderate ADRD and higher prevalence of mild cognitive impairment or suspected prodromal MND at baseline, but were more likely to have received their MND diagnosis from a specialist. After adjustment for potential baseline confounders via inverse probability of treatment weighting, all covariates were well-balanced between groups (absolute standardized differences <0.1).

**Table 1:**
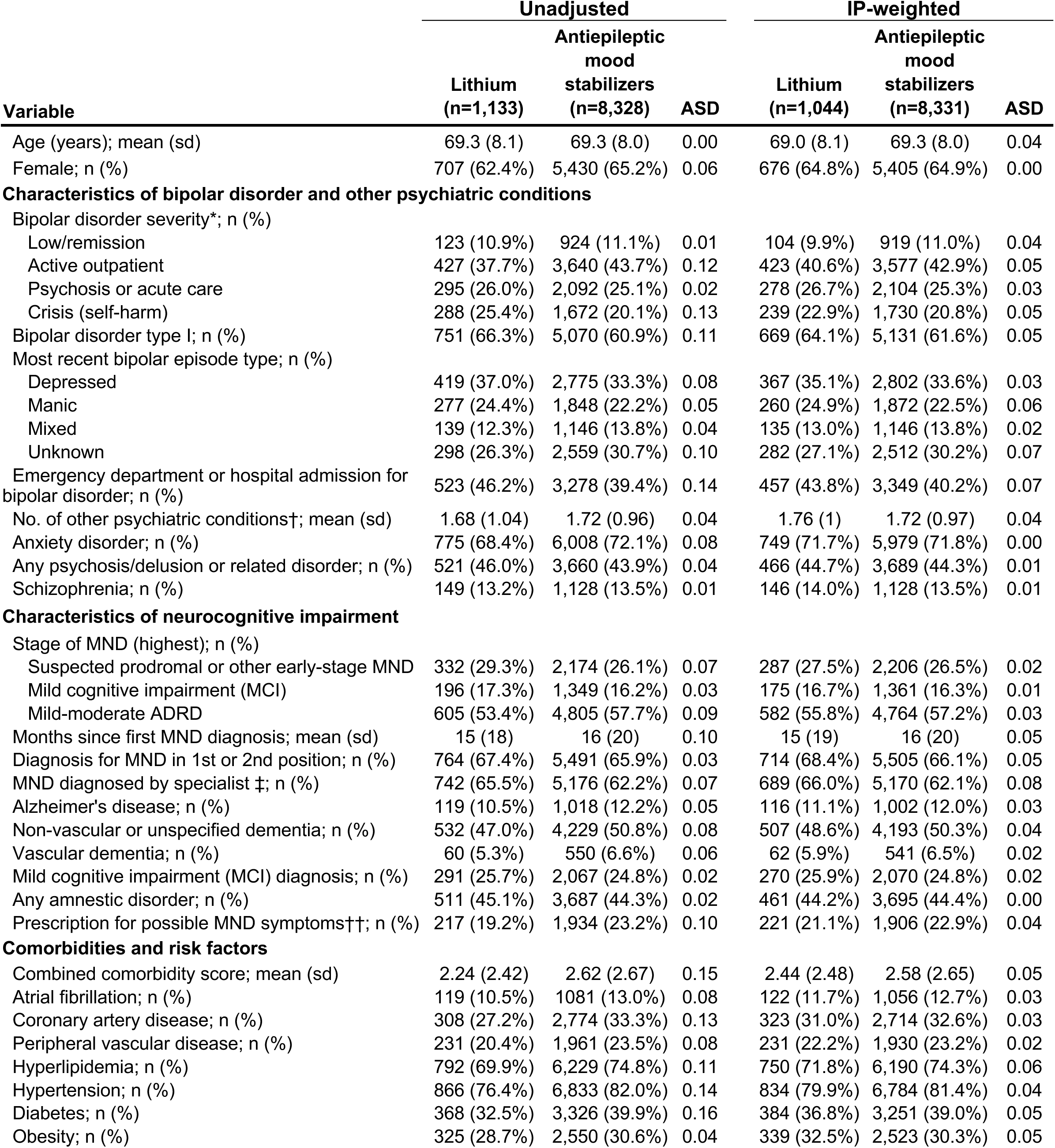
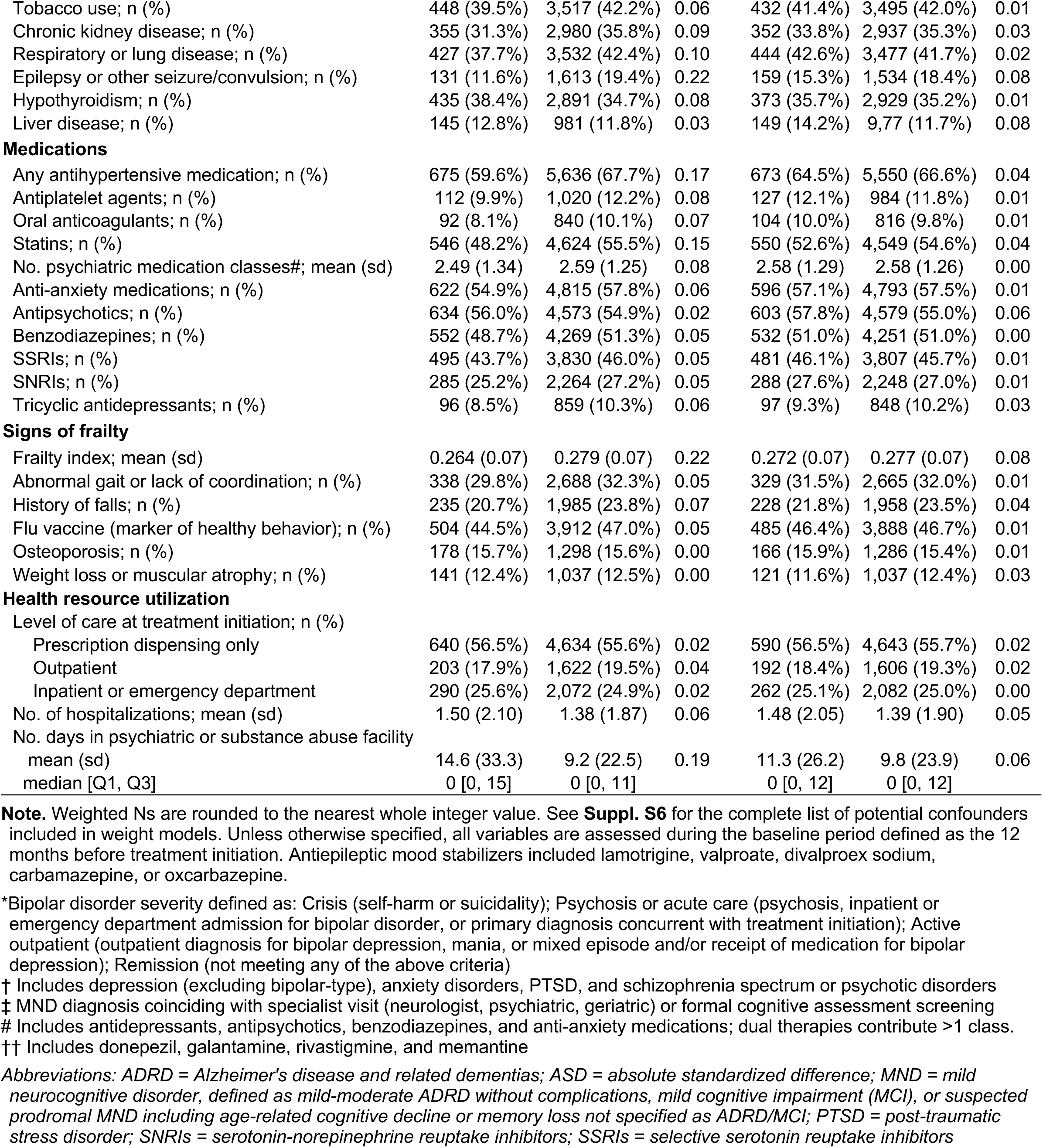
Baseline characteristics of Medicare beneficiaries included in the target trial emulation during the 1-year period before treatment initiation.

| Variable | Unadjusted |  |  | IP-weighted |  |  |
| --- | --- | --- | --- | --- | --- | --- |
|  | Lithium<br>(n=1,133) | Antiepileptic mood<br>stabilizers<br>(n=8,328) | ASD | Lithium<br>(n=1,044) | Antiepileptic mood<br>stabilizers<br>(n=8,331) | ASD |
| Age (years); mean (sd) | 69.3 (8.1) | 69.3 (8.0) | 0.00 | 69.0 (8.1) | 69.3 (8.0) | 0.04 |
| Female; n (%) | 707 (62.4%) | 5,430 (65.2%) | 0.06 | 676 (64.8%) | 5,405 (64.9%) | 0.00 |
| **Characteristics of bipolar disorder and other psychiatric conditions** |  |  |  |  |  |  |
| Bipolar disorder severity*; n (%) |  |  |  |  |  |  |
| Low/remission | 123 (10.9%) | 924 (11.1%) | 0.01 | 104 (9.9%) | 919 (11.0%) | 0.04 |
| Active outpatient | 427 (37.7%) | 3,640 (43.7%) | 0.12 | 423 (40.6%) | 3,577 (42.9%) | 0.05 |
| Psychosis or acute care | 295 (26.0%) | 2,092 (25.1%) | 0.02 | 278 (26.7%) | 2,104 (25.3%) | 0.03 |
| Crisis (self-harm) | 288 (25.4%) | 1,672 (20.1%) | 0.13 | 239 (22.9%) | 1,730 (20.8%) | 0.05 |
| Bipolar disorder type I; n (%) | 751 (66.3%) | 5,070 (60.9%) | 0.11 | 669 (64.1%) | 5,131 (61.6%) | 0.05 |
| Most recent bipolar episode type; n (%) |  |  |  |  |  |  |
| Depressed | 419 (37.0%) | 2,775 (33.3%) | 0.08 | 367 (35.1%) | 2,802 (33.6%) | 0.03 |
| Manic | 277 (24.4%) | 1,848 (22.2%) | 0.05 | 260 (24.9%) | 1,872 (22.5%) | 0.06 |
| Mixed | 139 (12.3%) | 1,146 (13.8%) | 0.04 | 135 (13.0%) | 1,146 (13.8%) | 0.02 |
| Unknown | 298 (26.3%) | 2,559 (30.7%) | 0.10 | 282 (27.1%) | 2,512 (30.2%) | 0.07 |
| Emergency department or hospital admission for bipolar disorder; n (%) | 523 (46.2%) | 3,278 (39.4%) | 0.14 | 457 (43.8%) | 3,349 (40.2%) | 0.07 |
| No. of other psychiatric conditions†; mean (sd) | 1.68 (1.04) | 1.72 (0.96) | 0.04 | 1.76 (1) | 1.72 (0.97) | 0.04 |
| Anxiety disorder; n (%) | 775 (68.4%) | 6,008 (72.1%) | 0.08 | 749 (71.7%) | 5,979 (71.8%) | 0.00 |
| Any psychosis/delusion or related disorder; n (%) | 521 (46.0%) | 3,660 (43.9%) | 0.04 | 466 (44.7%) | 3,689 (44.3%) | 0.01 |
| Schizophrenia; n (%) | 149 (13.2%) | 1,128 (13.5%) | 0.01 | 146 (14.0%) | 1,128 (13.5%) | 0.01 |
| **Characteristics of neurocognitive impairment** |  |  |  |  |  |  |
| Stage of MND (highest); n (%) |  |  |  |  |  |  |
| Suspected prodromal or other early-stage MND | 332 (29.3%) | 2,174 (26.1%) | 0.07 | 287 (27.5%) | 2,206 (26.5%) | 0.02 |
| Mild cognitive impairment (MCI) | 196 (17.3%) | 1,349 (16.2%) | 0.03 | 175 (16.7%) | 1,361 (16.3%) | 0.01 |
| Mild-moderate ADRD | 605 (53.4%) | 4,805 (57.7%) | 0.09 | 582 (55.8%) | 4,764 (57.2%) | 0.03 |
| Months since first MND diagnosis; mean (sd) | 15 (18) | 16 (20) | 0.10 | 15 (19) | 16 (20) | 0.05 |
| Diagnosis for MND in 1st or 2nd position; n (%) | 764 (67.4%) | 5,491 (65.9%) | 0.03 | 714 (68.4%) | 5,505 (66.1%) | 0.05 |
| MND diagnosed by specialist ‡; n (%) | 742 (65.5%) | 5,176 (62.2%) | 0.07 | 689 (66.0%) | 5,170 (62.1%) | 0.08 |
| Alzheimer's disease; n (%) | 119 (10.5%) | 1,018 (12.2%) | 0.05 | 116 (11.1%) | 1,002 (12.0%) | 0.03 |
| Non-vascular or unspecified dementia; n (%) | 532 (47.0%) | 4,229 (50.8%) | 0.08 | 507 (48.6%) | 4,193 (50.3%) | 0.04 |
| Vascular dementia; n (%) | 60 (5.3%) | 550 (6.6%) | 0.06 | 62 (5.9%) | 541 (6.5%) | 0.02 |
| Mild cognitive impairment (MCI) diagnosis; n (%) | 291 (25.7%) | 2,067 (24.8%) | 0.02 | 270 (25.9%) | 2,070 (24.8%) | 0.02 |
| Any amnesic disorder; n (%) | 511 (45.1%) | 3,687 (44.3%) | 0.02 | 461 (44.2%) | 3,695 (44.4%) | 0.00 |
| Prescription for possible MND symptoms††; n (%) | 217 (19.2%) | 1,934 (23.2%) | 0.10 | 221 (21.1%) | 1,906 (22.9%) | 0.04 |
| **Comorbidities and risk factors** |  |  |  |  |  |  |
| Combined comorbidity score; mean (sd) | 2.24 (2.42) | 2.62 (2.67) | 0.15 | 2.44 (2.48) | 2.58 (2.65) | 0.05 |
| Atrial fibrillation; n (%) | 119 (10.5%) | 1081 (13.0%) | 0.08 | 122 (11.7%) | 1,056 (12.7%) | 0.03 |
| Coronary artery disease; n (%) | 308 (27.2%) | 2,774 (33.3%) | 0.13 | 323 (31.0%) | 2,714 (32.6%) | 0.03 |
| Peripheral vascular disease; n (%) | 231 (20.4%) | 1,961 (23.5%) | 0.08 | 231 (22.2%) | 1,930 (23.2%) | 0.02 |
| Hyperlipidemia; n (%) | 792 (69.9%) | 6,229 (74.8%) | 0.11 | 750 (71.8%) | 6,190 (74.3%) | 0.06 |
| Hypertension; n (%) | 866 (76.4%) | 6,833 (82.0%) | 0.14 | 834 (79.9%) | 6,784 (81.4%) | 0.04 |
| Diabetes; n (%) | 368 (32.5%) | 3,326 (39.9%) | 0.16 | 384 (36.8%) | 3,251 (39.0%) | 0.05 |
| Obesity; n (%) | 325 (28.7%) | 2,550 (30.6%) | 0.04 | 339 (32.5%) | 2,523 (30.3%) | 0.05 |
| Tobacco use; n (%) | 448 (39.5%) | 3,517 (42.2%) | 0.06 | 432 (41.4%) | 3,495 (42.0%) | 0.01 |
| Chronic kidney disease; n (%) | 355 (31.3%) | 2,980 (35.8%) | 0.09 | 352 (33.8%) | 2,937 (35.3%) | 0.03 |
| Respiratory or lung disease; n (%) | 427 (37.7%) | 3,532 (42.4%) | 0.10 | 444 (42.6%) | 3,477 (41.7%) | 0.02 |
| Epilepsy or other seizure/convulsion; n (%) | 131 (11.6%) | 1,613 (19.4%) | 0.22 | 159 (15.3%) | 1,534 (18.4%) | 0.08 |
| Hypothyroidism; n (%) | 435 (38.4%) | 2,891 (34.7%) | 0.08 | 373 (35.7%) | 2,929 (35.2%) | 0.01 |
| Liver disease; n (%) | 145 (12.8%) | 981 (11.8%) | 0.03 | 149 (14.2%) | 9,77 (11.7%) | 0.08 |
| Medications |  |  |  |  |  |  |
| Any antihypertensive medication; n (%) | 675 (59.6%) | 5,636 (67.7%) | 0.17 | 673 (64.5%) | 5,550 (66.6%) | 0.04 |
| Antiplatelet agents; n (%) | 112 (9.9%) | 1,020 (12.2%) | 0.08 | 127 (12.1%) | 984 (11.8%) | 0.01 |
| Oral anticoagulants; n (%) | 92 (8.1%) | 840 (10.1%) | 0.07 | 104 (10.0%) | 816 (9.8%) | 0.01 |
| Statins; n (%) | 546 (48.2%) | 4,624 (55.5%) | 0.15 | 550 (52.6%) | 4,549 (54.6%) | 0.04 |
| No. psychiatric medication classes#; mean (sd) | 2.49 (1.34) | 2.59 (1.25) | 0.08 | 2.58 (1.29) | 2.58 (1.26) | 0.00 |
| Anti-anxiety medications; n (%) | 622 (54.9%) | 4,815 (57.8%) | 0.06 | 596 (57.1%) | 4,793 (57.5%) | 0.01 |
| Antipsychotics; n (%) | 634 (56.0%) | 4,573 (54.9%) | 0.02 | 603 (57.8%) | 4,579 (55.0%) | 0.06 |
| Benzodiazepines; n (%) | 552 (48.7%) | 4,269 (51.3%) | 0.05 | 532 (51.0%) | 4,251 (51.0%) | 0.00 |
| SSRIs; n (%) | 495 (43.7%) | 3,830 (46.0%) | 0.05 | 481 (46.1%) | 3,807 (45.7%) | 0.01 |
| SNRIs; n (%) | 285 (25.2%) | 2,264 (27.2%) | 0.05 | 288 (27.6%) | 2,248 (27.0%) | 0.01 |
| Tricyclic antidepressants; n (%) | 96 (8.5%) | 859 (10.3%) | 0.06 | 97 (9.3%) | 848 (10.2%) | 0.03 |
| Signs of frailty |  |  |  |  |  |  |
| Frailty index; mean (sd) | 0.264 (0.07) | 0.279 (0.07) | 0.22 | 0.272 (0.07) | 0.277 (0.07) | 0.08 |
| Abnormal gait or lack of coordination; n (%) | 338 (29.8%) | 2,688 (32.3%) | 0.05 | 329 (31.5%) | 2,665 (32.0%) | 0.01 |
| History of falls; n (%) | 235 (20.7%) | 1,985 (23.8%) | 0.07 | 228 (21.8%) | 1,958 (23.5%) | 0.04 |
| Flu vaccine (marker of healthy behavior); n (%) | 504 (44.5%) | 3,912 (47.0%) | 0.05 | 485 (46.4%) | 3,888 (46.7%) | 0.01 |
| Osteoporosis; n (%) | 178 (15.7%) | 1,298 (15.6%) | 0.00 | 166 (15.9%) | 1,286 (15.4%) | 0.01 |
| Weight loss or muscular atrophy; n (%) | 141 (12.4%) | 1,037 (12.5%) | 0.00 | 121 (11.6%) | 1,037 (12.4%) | 0.03 |
| Health resource utilization |  |  |  |  |  |  |
| Level of care at treatment initiation; n (%) |  |  |  |  |  |  |
| Prescription dispensing only | 640 (56.5%) | 4,634 (55.6%) | 0.02 | 590 (56.5%) | 4,643 (55.7%) | 0.02 |
| Outpatient | 203 (17.9%) | 1,622 (19.5%) | 0.04 | 192 (18.4%) | 1,606 (19.3%) | 0.02 |
| Inpatient or emergency department | 290 (25.6%) | 2,072 (24.9%) | 0.02 | 262 (25.1%) | 2,082 (25.0%) | 0.00 |
| No. of hospitalizations; mean (sd) | 1.50 (2.10) | 1.38 (1.87) | 0.06 | 1.48 (2.05) | 1.39 (1.90) | 0.05 |
| No. days in psychiatric or substance abuse facility |  |  |  |  |  |  |
| mean (sd) | 14.6 (33.3) | 9.2 (22.5) | 0.19 | 11.3 (26.2) | 9.8 (23.9) | 0.06 |
| median [Q1, Q3] | 0 [0, 15] | 0 [0, 11] |  | 0 [0, 12] | 0 [0, 12] |  |

**Note.** Weighted Ns are rounded to the nearest whole integer value. See **Suppl. S6** for the complete list of potential confounders included in weight models. Unless otherwise specified, all variables are assessed during the baseline period defined as the 12 months before treatment initiation. Antiepileptic mood stabilizers included lamotrigine, valproate, divalproex sodium, carbamazepine, or oxcarbazepine.
\*Bipolar disorder severity defined as: Crisis (self-harm or suicidality); Psychosis or acute care (psychosis, inpatient or emergency department admission for bipolar disorder, or primary diagnosis concurrent with treatment initiation); Active outpatient (outpatient diagnosis for bipolar depression, mania, or mixed episode and/or receipt of medication for bipolar depression); Remission (not meeting any of the above criteria)
† Includes depression (excluding bipolar-type), anxiety disorders, PTSD, and schizophrenia spectrum or psychotic disorders
‡ MND diagnosis coinciding with specialist visit (neurologist, psychiatric, geriatric) or formal cognitive assessment screening

Overall, 2,249 patients experienced incident progression to advanced ADRD over a median follow-up of 1.8 years (interquartile range, 0.7-3.5 years), and 697 patients had a long-term care stay with ADRD over a median follow-up of 2.2 years (interquartile range, 0.9-3.9 years). (**Suppl. S11**) The adjusted 5-year risk ratio for progression to advanced ADRD comparing initiation of lithium versus antiepileptic mood stabilizers was 0.87 (95% confidence interval 0.78, 0.99), the risk difference was -4.19% (-7.36%, -0.02%), and the hazard ratio was 0.85 (0.73, 0.98) (**Fig. 1A**). For long-term care stay with ADRD, the adjusted risk ratio was 0.75 (0.56, 0.97), the risk difference was -2.95% (-5.32%, -0.03%) and the hazard ratio was 0.70 (0.53, 0.91) (**Fig. 1B**).

**Fig. 1:**
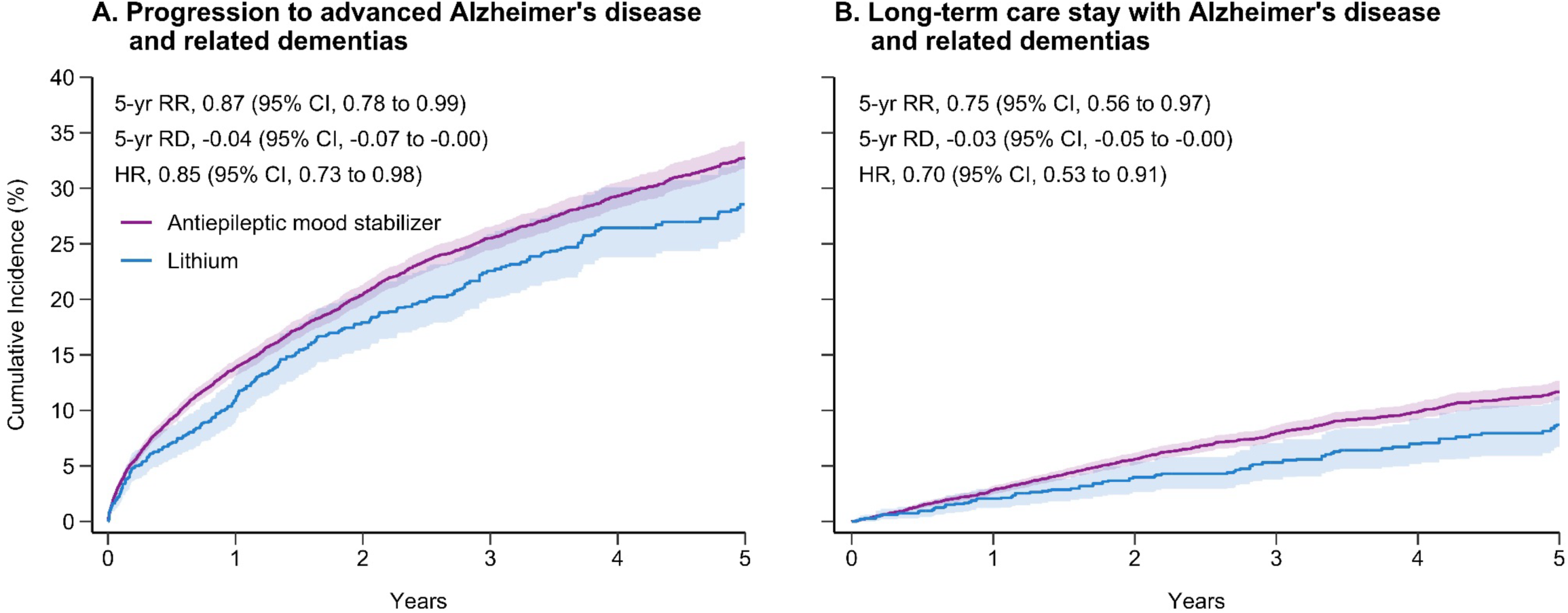
Adjusted 5-year risks of outcomes for progression of Alzheimer’s disease and related dementias in patients initiating lithium versus antiepileptic mood stabilizers (Medicare) **Note.** Risks were estimated with cumulative incidence functions and hazard ratios with Cox proportional hazard models. Inverse probability of treatment weighting was used to adjust for potential baseline confounding. Progression to advanced Alzheimer’s disease and related dementias (A) was defined as an incident diagnosis for severe or complicated Alzheimer’s disease or related dementias. Long-term care stay with Alzheimer’s disease and related dementias (B) was defined as a diagnosis for Alzheimer’s disease or related dementias overlapping with a continuous stay of at least 100 days in a nursing home or suspected long-term care facility. *HR = hazard ratio, RD = risk difference, RR = risk ratio.*

In subgroup analyses (**Fig 2**), risk reductions were most pronounced among patients with MCI or mild-moderate ADRD at the time of treatment start, with no difference for those with suspected prodromal disease. The risk of progression to advanced ADRD and long-term care stay with ADRD increased across the gradient of MND subgroups, which represent sequential stages along the continuum of ADRD progression.

**Fig. 2:**
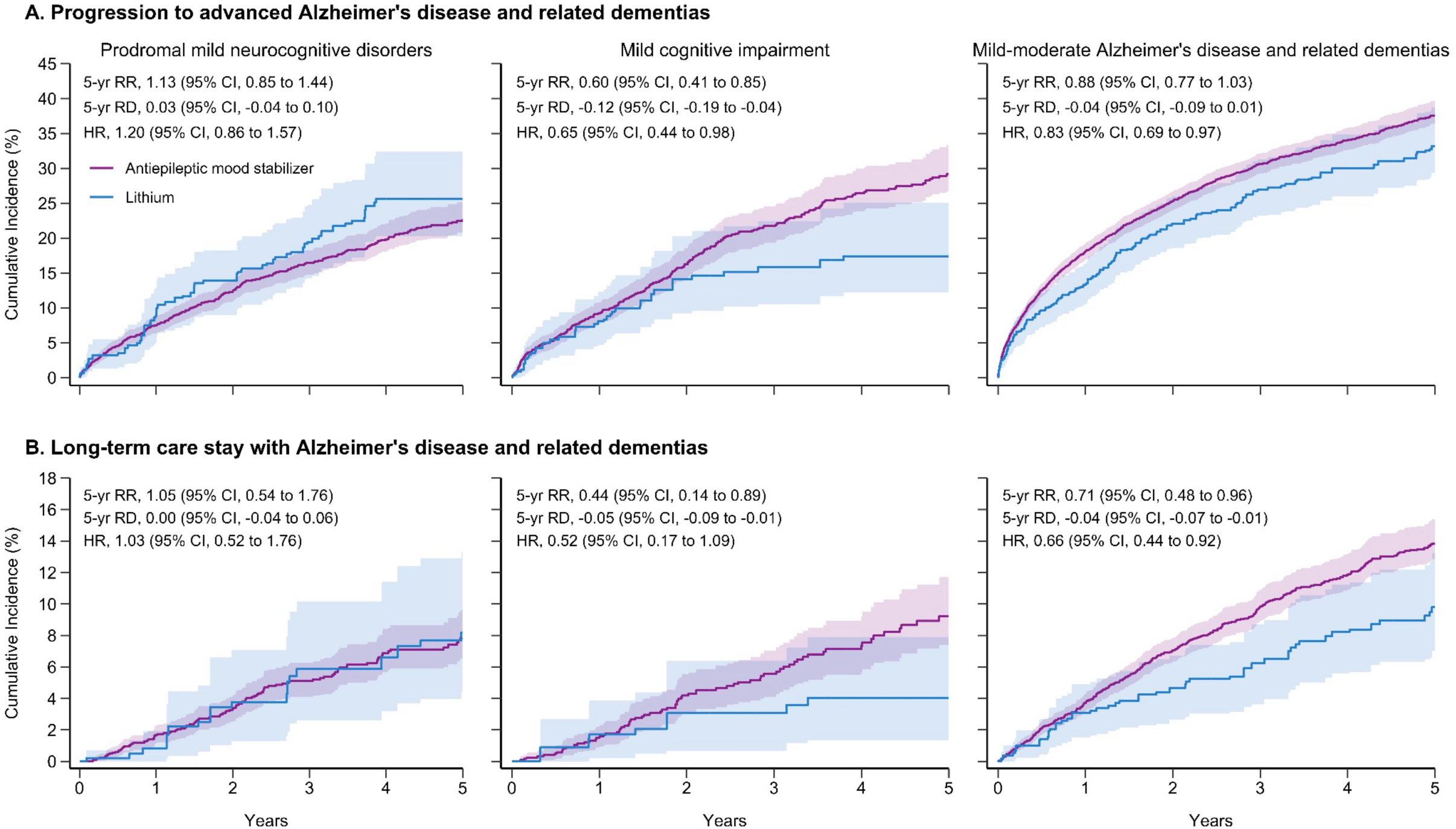
Adjusted 5-year risks of outcomes for progression of Alzheimer’s disease and related dementias in patients initiating lithium versus antiepileptic mood stabilizers, stratified by stage of mild neurocognitive disorder at baseline (Medicare) **Note.** Risks were estimated with cumulative incidence functions and hazard ratios with Cox proportional hazard models. Inverse probability of treatment weighting was used to adjust for potential baseline confounding. Subgroups for baseline stage of mild neurocognitive disorder were based on the most severe neurocognitive impairment diagnosis code recorded during the baseline period (suspected prodromal disease > mild cognitive impairment > mild-moderate Alzheimer’s disease and related dementias). Progression to advanced Alzheimer’s disease and related dementias (A) was defined as an incident diagnosis for severe or complicated Alzheimer’s disease and related dementias. Long-term care stay with Alzheimer’s disease and related dementias (B) was defined as a diagnosis for Alzheimer’s disease and related dementias overlapping with a continuous stay of at least 100 days in a nursing home or suspected long-term care facility. *HR = hazard ratio, RD = risk difference, RR = risk ratio*.

Emulating the target trial in two commercial claims databases yielded congruent findings. The adjusted 5-year risk ratio for progression to advanced ADRD was 0.90 (0.68,1.14) in Optum CDM and 0.57 (0.30, 0.90) in Merative MarketScan. Risk ratios for long-term care stay with ADRD were similar but had wide confidence intervals (**Suppl. S8**). Fixed-effect meta-analysis pooling estimates across all three claims databases yielded risk ratios of 0.84 (0.77, 0.92) for progression to advanced ADRD and 0.74 (0.60, 0.90) for long-term care stay with ADRD (**Table S12**).

### Sensitivity Analyses

We conducted nine pre-specified sensitivity analyses (SAs) to assess robustness (**Fig. 3**, **Suppl. S10**). Results were directionally consistent with findings from our main analyses across:

- <u>Alternative eligibility criteria</u>: excluding persons with other possible causes of dementia (SA1), excluding those with diagnoses for non-bipolar antiepileptic indications (SA2), requiring more stringent definitions for bipolar disorder (SA3) and MND (SA4)
- <u>Accounting for competing events</u>: composite outcome of ADRD progression *or* death to account for death as a potential competing event (SA5)
- <u>Alternative design parameters</u>: shortening the treatment washout period from 365 to 180 days to reflect real-world treatment patterns (SA6)
- <u>Alternative outcome definitions intended to mitigate misclassification</u>: Advanced ADRD diagnosis in primary position (SA7), advanced ADRD diagnosis as admitting/primary diagnosis in inpatient or long-term care setting (SA8), requiring an incident prescription for cholinesterase inhibitors or memantine following diagnosis for advanced ADRD (SA9).^17^

**Fig. 3:**
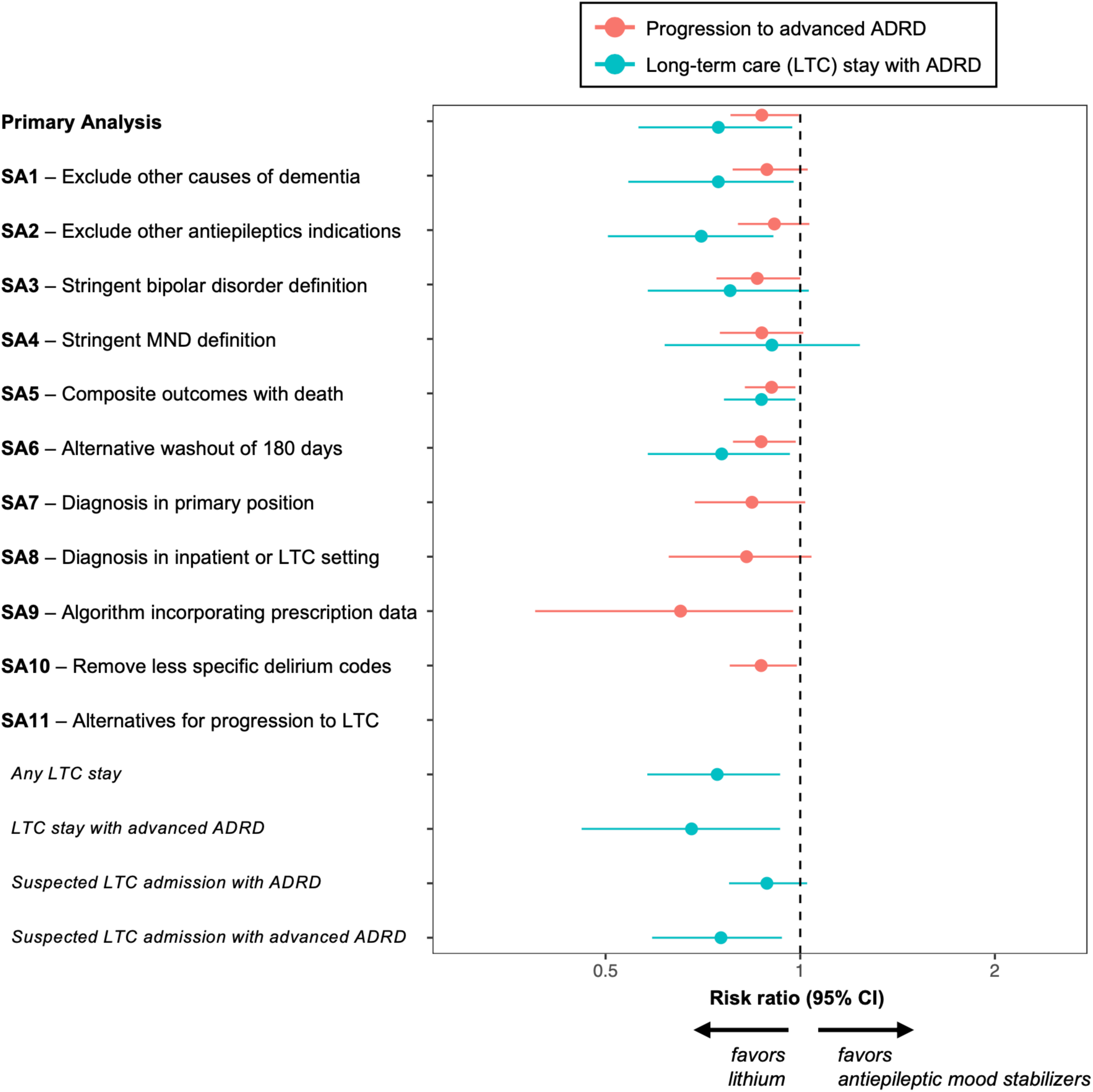
Adjusted 5-year risk ratio estimates from sensitivity analyses (Medicare) **Note**. SA1-SA9 were pre-specified, while SA10-SA11 were pursued as post-hoc robustness checks. *ADRD = Alzheimer’s disease and related dementia, AMS = antiepileptic mood stabilizers, LTC = long-term care, SA = sensitivity analysis*

Two post-hoc sensitivity analyses also confirmed main findings. Lithium initiators had lower risk of progression to advanced ADRD when excluding codes for dementia with delirium (SA10)^17^ and across alternative definitions of the long-term care admission-based outcome (SA11). Extended results for additional outcomes and subgroup analyses are available in the **Supplementary Materials**.

### Validations in information-rich subsets of Medicare beneficiaries

Use of claims data to study neurocognitive diseases is challenging due to incomplete measurement of cognitive and functional impairments in data collected primarily for billing purposes.^18^ In our study, the potential for misclassification and unmeasured confounding is potentially amplified by the overlapping indications and sequelae of psychiatric and neurodegenerative diseases (see **Extended Table 2**). To understand the potential for bias, we conducted three validations leveraging detailed cognitive and functional status information available for a subset of Medicare beneficiaries. These exercises addressed three critical questions related to potential information and confounding bias in claims data: (Q1) whether eligibility criteria adequately identified MND while excluding advanced ADRD; (Q2) whether residual confounding in cognitive status persisted after adjustment for covariates measured in claims data; and (Q3) whether claims-based criteria for ADRD progression aligned with other validated instruments of impairment severity.

### Q1: Do claims-based eligibility criteria adequately identify mild neurocognitive disorder while excluding advanced ADRD?

We assessed cognitive status at baseline using two supplementary sources: the Patient Assessment Instrument (PAI) administered in inpatient rehabilitation facilities, and the Outcome and Assessment Information Set (OASIS) administered during home health visits. Items from these sources have been validated^19,20^ against the Mini-Mental State Examination and other gold standard instruments and are used for ADRD identification in other studies.^21,22^

Our target trial was designed to include patients across the early-to-middle MND/ADRD continuum at risk for progression to advanced disease. To evaluate whether eligibility criteria adequately identified MND while excluding advanced ADRD, we applied algorithms for severe dementia^21^ using IRF-PAI and OASIS instruments (Methods). Among 2,036 patients with baseline assessments, only 11 (0.5%) met criteria for severe dementia (**Fig. 4A, Table S13**), supporting the reliability of diagnosis-based eligibility criteria.

**Fig. 4:**
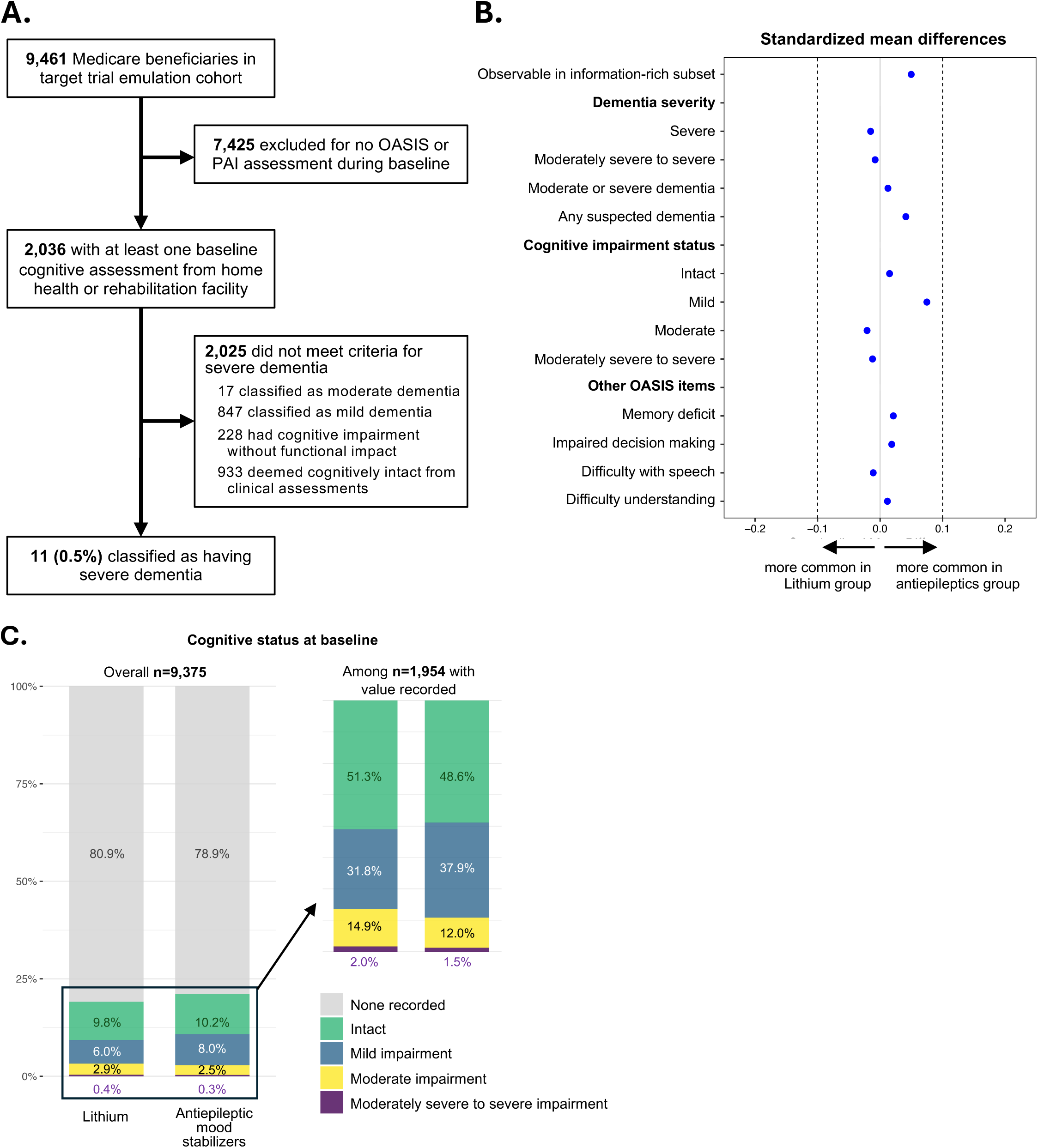
Pre-treatment cognitive function information from an information-rich subset of Medicare beneficiaries **Note.** Variables are extracted from the most recent assessment during the baseline period, defined as the 12 months before treatment initiation. Assessments include the Outcome and Assessment Information Set (OASIS) and Patient Assessment Instrument (PAI). **A.** Flow diagram for population identified in validation Q1. Severe dementia was defined using OASIS and PAI data as severe cognitive impairment plus three impaired daily living activities and dependency in at least one advanced daily living activity; see Methods section for details. **B.** Standardized mean differences comparing balance for OASIS/PAI items between treatment groups. Standardized mean differences were calculated using the overall weighted population (n=1,044 lithium, n=8,331 antiepileptic mood stabilizers). **C.** Distribution of baseline cognitive status stratified by treatment group. Percentages are shown for the overall weighted population and among the subset with at least one assessment record (see inset). Cognitive status is derived from the OASIS Cognitive Function instrument and IRF-PAI modified Cognitive Function Scale (Methods).

### Q2: Is there residual confounding in baseline cognitive status across treatment groups?

Using the same baseline data as validation Q1, we examined whether lithium and antiepileptic mood stabilizer initiators differed on detailed cognitive measures after adjustment for other potential confounders. Baseline cognitive function and other PAI/OASIS indicators of impairment were well-balanced between treatment groups after inverse probability weighting **(Fig. 4B-4C, Table S13**), strengthening the plausibility of baseline equipoise in cognitive status.

### Q3: Does claims-based ADRD progression during follow-up agree with impairment severity measured in nursing facility assessments?

A key concern for claims-based studies is outcome misclassification. While broader ADRD diagnostic codes have been validated,^17,23,24^ specific sub-codes distinguishing clinical stages of disease progression have not. To address this concern, we assessed whether our claims-based progression measures correlated with validated instruments for cognitive and functional impairment from the Minimum Data Set.^25–29^ These assessments confirmed strong concordance between claims-based progression stages and measured impairment severity.

The Minimum Data Set, a standardized assessment tool for nursing facility residents, was available for 2,285 patients from the target trial emulation who were admitted to a nursing facility during follow-up (**Fig. 5A**). We extracted and compared Minimum Data Set items for cognitive function and self-performance of activities of daily living across our four claims-based stages of neurocognitive disorder (suspected prodromal MND, MCI without ADRD, mild-moderate ADRD, and advanced ADRD) (**Fig. 5B-5D, Table S13**).

**Fig. 5:**
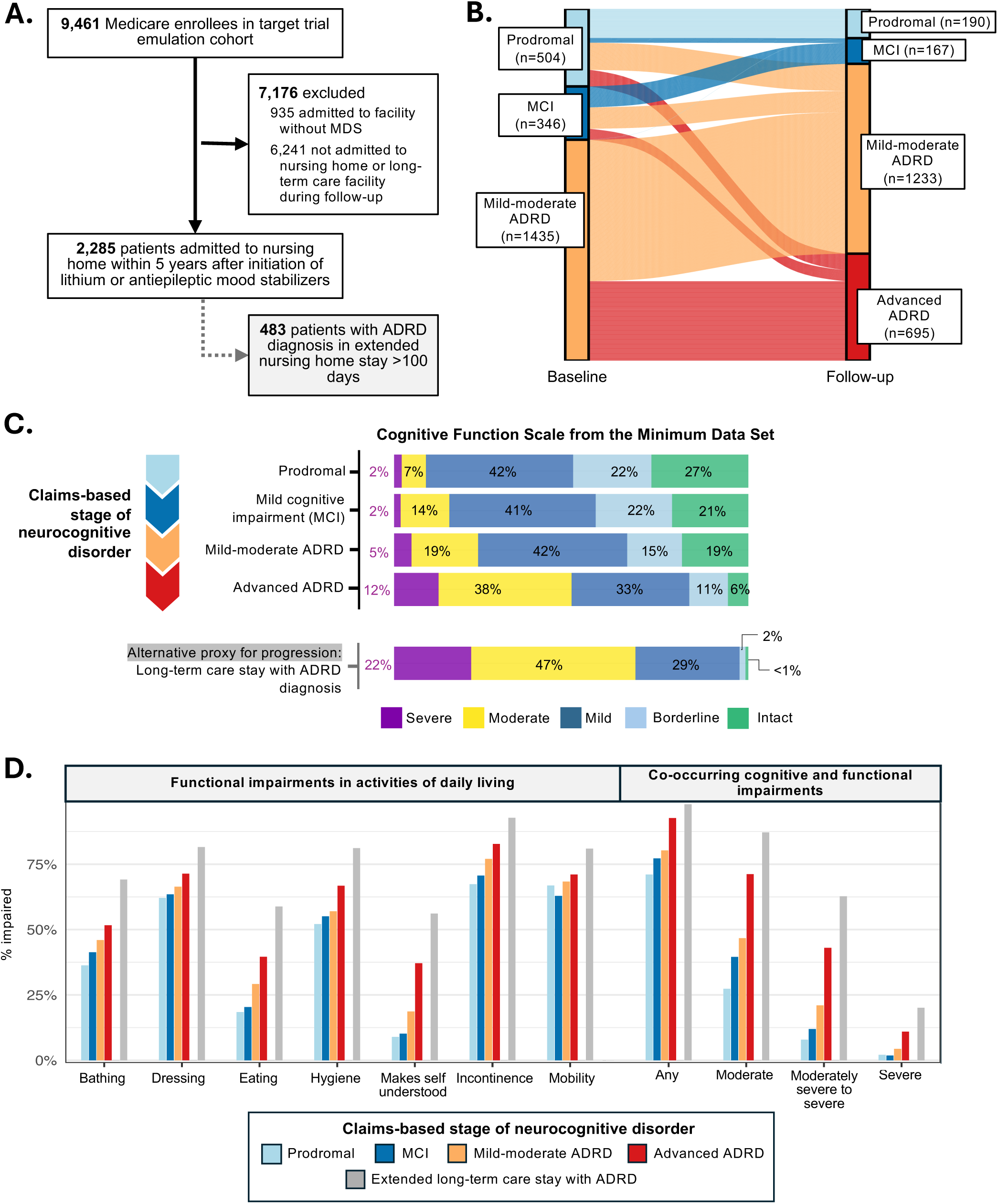
Agreement between claims-based stage of neurocognitive disorder during follow-up and cognitive and functional information from an information-rich subset of Medicare beneficiaries **A.** Flow diagram for population identified in validation Q3, comprising n=2,285 patients admitted to a nursing facility during follow-up. Data are from the first (incident) admission. **B.** Sankey diagram of claims-based neurocognitive disorder trajectories for n=2,285 patients from baseline (365-day period before treatment initiation) to follow-up (incident admission to nursing facility within 5-years after treatment initiation). Patients were assigned a claims-based stage according to the most severe neurocognitive impairment diagnosis code recorded through each time period (prodromal MND < MCI < mild-moderate ADRD < advanced ADRD). **C.** Cognitive Function Scale from linked Minimum Data Set, stratified by claims-based impairment stage at admission, and shown separately for patients meeting the second main outcome of a long-term care stay with ADRD. **D**. Proportion of patients with Minimum Data Set-documented impairments in activities of daily living, stratified by claims-based impairment stage at admission. The right panel shows the proportion of patients with co-occurring cognitive and functional impairments, defined to emulate stages from the Alzheimer’s Global Deterioration Scale (see Methods). *Abbreviations: ADRD = Alzheimer’s disease and related dementias; MDS = Minimum Data Set v3.0; MCI = mild cognitive impairment; MND = mild neurocognitive disorder*.

Figure 5B shows claims-based neurocognitive disorder trajectories for this population from baseline to the time of the incident nursing facility admission during follow-up. At admission, claims-based disease staging classified 30.4% as having progressed to advanced ADRD, 54.0% as mild-moderate ADRD, 7.3% as MCI, and 8.3% as suspected prodromal MND (Fig. 5B). Across all Minimum Data Set measures, patients meeting our claims-based definition of advanced ADRD had the greatest cognitive and functional impairment (Fig. 5C**–5D**).

Moreover, as claims stage increased from suspected prodromal MND through advanced ADRD, the distribution of ratings shifted monotonically toward greater impairment, supporting the use of claims-based stages as an ordinal severity gradient. For example, the proportion of patients with moderate to severe cognitive and functional impairment increased from 9% in the suspected prodromal MND group to 16% for MCI, 47% for mild-moderate ADRD, and 71% for advanced ADRD (Fig. 5D, **Table S13**). Claims-based stage of neurocognitive disorder also correlated with nursing facility length of stay (mean 71 days for suspected prodromal MND, 83 days for MCI, 109 days for mild-moderate ADRD, and 154 days for advanced ADRD). Similar monotonic trends were observed for impairments in activities of daily living, with advanced ADRD patients more likely to require assistance with bathing, dressing, eating, and other tasks (Fig. 5D**, Table S13).**

Impairment severity was even more pronounced for patients experiencing the outcome of a long-term care stay (≥100 days) with ADRD. Among the 483 patients with this outcome during follow-up who had corresponding Minimum Data Set assessments, 87% had moderate to severe impairments in both cognitive and functional measures (Fig. 5C**-5D**, Table S14).

## DISCUSSION

Across three nationwide US claims databases, among patients with bipolar disorder and MND, initiation of lithium was associated with a lower risk of ADRD progression compared to initiation of antiepileptic mood stabilizers. These findings were consistent across a range of outcomes and sensitivity analyses.

The potential role of lithium in preventing ADRD onset or progression has been extensively debated. While two cohort studies have found lower risk of incident dementia with long-term lithium use,^5,6^ others have not.^7–9,30^ Many population-based studies were susceptible to confounding by indication or immortal time bias,^31^ often stemming from problematic design choices such as comparing lithium users to untreated controls.

Evidence for lithium’s effects on disease progression among patients with pre-existing cognitive impairment is limited to small randomized trials, typically with fewer than 100 participants. A recent meta-analysis^4^ of six such trials found no significant difference between lithium and placebo on cognitive function scores (standardized mean difference of -0.21 [-0.49, 0.07]). Notably, four of the six contributing trials evaluated sub-therapeutic doses of conventional lithium salts (target range <0.6 mmol/L), which may be insufficient to achieve neuroprotective effects observed in preclinical models.

Our study design specifically addresses key limitations of prior research. First, using antiepileptic mood stabilizers as an active comparator reduces confounding by indication, as both drug classes are prescribed for the same underlying condition (bipolar disorder). Second, restricting to older adults with bipolar disorder and pre-existing MND reduces risks of outcome misclassification and etiologically implausible exposure windows – biases that may have affected prior studies that included adults of all ages and all levels of cognitive function. Third, by explicitly emulating a target trial^12^ with an intention-to-treat contrast, we avoid biases due to selection and immortal time, which may have afflicted other studies on this topic (e.g., those with duration-based exposure definitions^5,6,9,30^). Fourth, validations using detailed cognitive assessments provided reassurance against key threats to validity, namely residual confounding by baseline cognition and outcome misclassification. Finally, our population received therapeutic lithium doses for mood stabilization,^32^ substantially higher than the sub-therapeutic doses used in most dementia trials.

A recent study by Aron et al^1^ identified lithium orotate as the most promising lithium-containing salt for slowing ADRD progression, due to its low affinity for amyloid, which may reduce lithium sequestration by amyloid deposits. In contrast, lithium carbonate and citrate, the compounds studied here, may be more vulnerable to sequestration by amyloid and thus potentially less effective. However, the therapeutic doses used in routine bipolar disorder care^32^ may be sufficiently high to overcome partial sequestration, which could explain why our study found lower risks with lithium initiation while some trials using sub-therapeutic doses of conventional lithium salts did not.^4,33^ While therapeutic doses of lithium carbonate and citrate have important safety considerations (e.g., nephropathy^34^) our findings complement evidence from Aron et al, lending support to the broader hypothesis that lithium supplementation may slow cognitive decline. These converging lines of evidence support evaluation of optimized lithium formulations with improved brain bioavailability and reduced toxicity in adequately powered randomized trials.

While our rigorous design mitigates common limitations of prior studies, some caution in interpretation is warranted due to inherent limitations of lithium use patterns in routine care and challenges in administrative data capture. Key threats to validity and accompanying mitigation strategies are detailed in **Extended Data Table 2.**

For instance, clinical caution with lithium in severe kidney or cardiovascular disease may lead to preferential antiepileptic mood stabilizer use among medically complex patients or after lithium intolerance. Additionally, some antiepileptic mood stabilizer initiators may be treated for non-bipolar conditions (e.g., epilepsy, neuropathic pain) or may receive antiepileptics to address both bipolar disorder and these comorbidities. However, these factors were well-balanced after adjustment, and findings were consistent when excluding patients with non-bipolar antiepileptic mood stabilizer indications and using more stringent bipolar disorder criteria. Off-label antiepileptic use for dementia symptoms is also possible but would require substantial misclassification of both treatment indication and baseline cognitive status; validations found no evidence of either, and our exclusion of baseline nursing facility utilization minimizes this risk.^35^ Conversely, lithium may be preferentially prescribed for more severe bipolar disorder. Because bipolar disorder severity is a risk factor for ADRD,^36^ this may bias results conservatively, attenuating any potential protective effect of lithium relative to antiepileptic mood stabilizers.

Next, claims-based misclassification of MND and ADRD progression is also possible. Validation studies have found positive predictive values of 65-84% for claims-based dementia identification in Medicare,^17,23,24^ but none have assessed measurement performance in populations with psychiatric disease or validated codes distinguishing complicated from uncomplicated dementia. Our outcome definitions for ADRD progression may have higher specificity than typical incident dementia algorithms. For instance, the outcome for progression to advanced ADRD requires diagnosis of both dementia *and* documented complications, which may necessitate more thorough adjudication. Frequent healthcare interactions among persons with bipolar disorder may also improve longitudinal tracking of neurodegeneration relative to general populations used for diagnostic validation studies. Moreover, results remained consistent when using alternative definitions expected to improve specificity,^17,37^ including more stringent algorithms for MND eligibility criteria, refined definitions for ADRD progression, and additional outcomes incorporating long-term care admissions.

Our validation results provide additional support for claims-based measures of ADRD progression, though formal validation studies are needed. While our staging approach shares conceptual similarities with clinical diagnostic criteria^13,14^ it is necessarily limited to the information available in claims. We could not use biomarkers to confirm Alzheimer’s pathophysiology or measure impairments with validated instruments across the full cohort. Nonetheless, validations showed strong agreement between claims-based stages and detailed cognitive assessments within the information rich subset of Medicare beneficiaries. To our knowledge, this study is among the first attempts to use claims-based diagnoses to distinguish different phases of ADRD progression. This preliminary work opens new opportunities for leveraging Medicare claims to study ADRD natural history and potential disease-modifying interventions.

Several additional limitations would be expected to attenuate any potential protective effect of lithium, further strengthening confidence in our findings. For instance, our intention-to-treat approach may underestimate sustained treatment effects due to discontinuation or switching of medications during the 5-year follow-up period (average lithium treatment duration 1.2 years; **Suppl. S11**), and prior treatment with study medications before Medicare enrollment may attenuate observed differences between groups. Heterogeneity in dementia subtypes within our outcome - including vascular and other non-Alzheimer’s dementias - may also attenuate potential effects if lithium primarily targets amyloid-related pathology. Finally, the prodromal MND category, intended to capture early-stage disease before formal diagnosis of MCI/ADRD,^38^ may include some patients with deficits unrelated to ADRD pathophysiology or without objectively confirmed impairment. Subgroup analyses revealed a clear gradient in progression risk across MND categories, with the strongest lithium effects in the MCI and mild-moderate ADRD groups, suggesting that inclusion of less specific prodromal cases may dilute overall effects.

In conclusion, our study provides robust real-world data supporting lithium’s potential to slow ADRD progression in older adults with bipolar disorder. Randomized trials of safer, optimized lithium formulations are needed to confirm clinical benefit.

## Methods

### Protocol and design of the target trial and the observational emulations

We emulated a new-user, active comparator target trial comparing progression of Alzheimer’s disease and related dementias (ADRD) in patients initiating lithium versus antiepileptic mood stabilizers, among adults ≥55 years with bipolar disorder and a mild neurocognitive disorder (MND). We used three deidentified claims databases from the United States: Medicare, Merative MarketScan, and Optum Clinformatics Data Mart. The Medicare emulation was designated a priori as the main analysis given more reliable long-term follow-up of older adults, with Optum Clinformatics Data Mart and Merative MarketScan emulations serving as secondary replication analyses. The study design and analyses were pre-registered before accessing Medicare data, with only minor adaptations to pre-specified plans (**Table S2**).

**Extended Data Table 1** outlines key components of the target trial and its observational emulation in the claims databases. **Extended Data** Fig. 1 summarizes the study design.

### Data Sources

#### Medicare

We used claims data from the Centers for Medicare & Medicaid Services for a random 20% sample of Medicare beneficiaries from January 1, 2013 through December 31, 2022. Medicare is the primary U.S. health insurance program for adults aged ≥65 years and those under 65 with qualifying disabilities. We used research identifiable files with fee-for-service claims from from Medicare Parts A (institutional and hospital care), B (non-institutional services), and D (prescription drugs). Dates of death from the Master Beneficiary Summary File were linked via unique patient identifiers with specific service dates. An information-rich subset of beneficiaries also had linked assessment data from nursing facilities (Minimum Data Set v3.0), home health agencies (Outcome and Assessment Information Set), or inpatient rehabilitation facilities (Patient Assessment Instrument). These standardized instruments collect clinical, functional, and cognitive assessments at regular intervals or upon admission and discharge. Linked assessments were used for validation exercises and select eligibility criteria only (described below in detail).

#### Merative MarketScan

We used data from both the Merative MarketScan Commercial Claims and Encounters (CCAE) database and the Medicare Supplemental (MDCR) database, with claims from December 31, 2012 to December 31, 2022. The CCAE database contains medical and pharmacy claims data for individuals not eligible for Medicare, including active employees, dependents with employer-provided coverage, early retirees, and COBRA continuees. The MDCR database includes claims for Medicare-eligible individuals (and dependents) with Medicare Advantage or employer-sponsored Medicare supplemental plans. Both databases include claims from fee-for-service and capitated plans, covering inpatient services, outpatient services, and prescription drug events. Date of death records are available for a subset of deaths occurring within inpatient settings or via linkage to the Social Security Administration Death Master File.

#### Optum Clinformatics Data Mart

The Optum Clinformatics Data Mart database includes administrative medical and pharmacy claims from January 1, 2004 to May 31, 2025 for individuals enrolled in plans affiliated with a large national managed care organization. The database includes patients with complete medical and prescription drug coverage from employer-sponsored coverage, private health insurance, or Medicare Advantage plans. In addition to medical and pharmacy claims, date of death records are included via linkage with several external sources, including the Social Security Administration Death Master File and Medicare Beneficiary Summary File.

### Specification of the target trial

#### Eligibility

Patients would be eligible for inclusion in the target trial if they were ≥55 years old and, in the prior 12 months: had received a diagnosis of bipolar disorder, had received a diagnosis for a MND (defined as suspected prodromal MND including age-related cognitive decline or memory loss, mild cognitive impairment (MCI), or non-severe ADRD without complications); had received no prescriptions for lithium (lithium carbonate or citrate) or the selected antiepileptic mood stabilizers (valproate, divalproex, lamotrigine, carbamazepine, oxcarbazepine); had no Parkinson’s disease diagnosis; and had no record of care in a nursing facility or suspected long-term care setting. Patients with any history of advanced (complicated or severe) ADRD or with no information on their age and sex would be excluded.

#### Treatment strategies and assignment

Eligible patients would be randomly assigned to initiate lithium (lithium carbonate or citrate) or an antiepileptic mood stabilizer (lamotrigine, valproate, divalproex sodium, carbamazepine, or oxcarbazepine).

#### Outcomes

The main outcomes of interest would be (i) progression to advanced ADRD and (ii) a long-term care stay (at least 100 days) with ADRD.

#### Follow-up

Follow-up would start on the day patients met eligibility criteria and were assigned to initiate lithium (carbonate or citrate) or an antiepileptic mood stabilizer, respectively, and would end on the earliest occurrence of the outcome of interest, loss-to-follow-up, 5 years, administrative end of follow-up, or death.

#### Causal contrasts

The target trial would estimate intention-to-treat effects for each outcome, comparing initiation of lithium with initiation of an antiepileptic mood stabilizer.

#### Statistical analysis

Intention-to-treat effects would be estimated as 5-year risk ratios and risk differences using cumulative incidence functions, considering death as a competing event. Hazard ratios would be estimated from a Cox proportional hazards model. Subgroup analyses would be conducted by MND stage at baseline (suspected prodromal MND not specified as ADRD or MCI, MCI without ADRD, mild-moderate ADRD), sex (female, male), and age (55–69, ≥70 years). Percentile-based 95% confidence intervals for risks, 5-year risk ratios and risk differences, and hazard ratios would be obtained from 1,000 non-parametric bootstrap samples.

### Emulation of the target trial

#### Eligibility

We used the same eligibility criteria as the target trial in the observational emulations, with operationalization of eligibility criteria in medical and pharmacy claims data.

#### Mild neurocognitive disorders

Alzheimer’s disease and related dementias (ADRD) are neurodegenerative conditions characterized by progressive cognitive and functional decline that ultimately interferes with daily living. The clinical continuum of ADRD can be categorized into preclinical or prodromal disease, involving asymptomatic pathology or subtle cognitive decline; MCI, characterized by changes in memory or cognition that are noticeable but do not interfere with daily life; and established dementia, when cognitive and functional impairments interfere with the ability to function.^14,39^ Dementia (i.e., ADRD) can be further separated into mild, moderate, and severe dementia, based on progressive cognitive and functional impairments that worsen over time. The target trial and observational emulation was designed to include patients across the early-to-middle stages of the ADRD continuum at risk for progression to advanced ADRD, which we refer to as mild neurocognitive disorders (MND).^13^ MND eligibility criteria were implemented with International Classification of Diseases (ICD) 9th and 10th revisions diagnosis codes for age-related cognitive decline or memory loss (termed "prodromal MND", as a proxy for suspected early-stage disease), MCI, and mild-to-moderate ADRD without complications (see **Suppl. S4**). The criterion for no prior diagnosis of advanced (complicated or severe) ADRD was enforced from the earliest start of available data through the date of treatment initiation (time zero). For the emulation in Medicare, we additionally excluded those with a Cognitive Functioning item (M1700) of 4 for “totally dependent state” from home health assessments during the 12-month baseline period (an additional proxy for advanced ADRD). The outcome of progression to advanced ADRD therefore represents disease progression beyond the baseline MND stage.

#### Other eligibility criteria

To ensure sufficient observability of eligibility criteria and other study variables, we required continuous enrollment for at least 12 months prior to treatment initiation (in CMS Medicare parts A, B, and D; in Merative MarketScan medical and pharmacy benefit; or in Optum CDM medical and pharmacy benefit). In addition to enforcing continuous enrollment in Medicare Parts A/B/D, we excluded patients with HMO/Part C enrollment, since complete observability of medical and pharmacy events cannot be guaranteed for persons enrolled in Medicare Advantage plans.

#### Treatment strategies and assignment

Among eligible patients, we identified those receiving a prescription for either lithium or one of the select antiepileptic mood stabilizers. Patients were therefore assigned treatment strategies consistent with their observed pharmacy claims data, and assignment was assumed to be approximately randomized conditional on measured confounders (**Suppl. S6**).

#### Outcomes

The outcomes were identical to those in the target trial and ascertained via medical claims from institutional and non-institutional settings using ICD-9 and ICD-10 diagnosis codes (see **Suppl. S4** for codes). Progression to advanced ADRD was operationalized as an incident diagnosis for severe or complicated ADRD. Long-term care stay with ADRD was operationalized as a diagnosis for ADRD overlapping with a contiguous stay of at least 100 days in a suspected long-term care setting (ascertained via place of service codes and other facility identifiers; see **Suppl. S4**).

#### Follow-up

Follow-up began at initiation of lithium or an antiepileptic mood stabilizer (time zero) and ended on the earliest of outcome occurrence, disenrollment leading to loss to follow-up, 5 years, administrative end of follow-up (Medicare: December 31, 2022; Merative MarketScan: December 31, 2023; Optum Clinformatics Data Mart: May 31, 2025), or death.

#### Causal contrasts and statistical analysis

Because treatment assignment was not randomized in the claims databases, we estimated an observational analogue of the intention-to-treat effect with adjustment for potential baseline confounding. For each outcome, we estimated cumulative incidences (risks) for each treatment strategy using non-parametric cumulative incidence functions, weighted by stabilized inverse probability of treatment weights (IPTWs). 5-year risk differences and risk ratios were calculated from these risks. Hazard ratios were estimated with Cox proportional hazard models, weighted by the same set of IPTWs. For all estimates, 95% CIs were computed using non-parametric bootstrapping across 1,000 samples.

Stabilized IPTWs were calculated as the marginal probability of initiating the treatment the patient actually received divided by the patient’s probability of initiating that treatment conditional on potential baseline confounders. Conditional treatment probabilities were obtained from a logistic regression model for lithium initiation that included potential baseline confounders, selected a priori as assumed common causes of initiating lithium or select antiepileptic mood stabilizers and ADRD progression. We assessed balance between treatment groups across >300 potential baseline confounders and selected a parsimonious subset for the final model to obtain absolute standardized differences of <0.2 for all potential confounders and ≤10 model degrees of freedom per exposed patient. IPTWs were truncated at the 99^th^ percentile to avoid undue influence by extreme weights. Distributions of estimated propensity scores and IPTWs are shown in **Suppl. S11.** In the main analyses of the observational emulation using the Medicare database, the following potential baseline confounders were included in the final propensity score model: age, sex, Medicaid dual eligibility, race, geographic region, combined comorbidity score,^40^ frailty index^41^ (modeled using a natural cubic spline with 3 degrees of freedom), atrial fibrillation, chronic kidney disease, chronic lung disease, diabetes, epilepsy or other convulsions, log number of epilepsy/convulsion events during baseline, diabetic neuropathy and related neuropathies, peripheral vascular disease, stroke or transient ischemic attack, coronary artery or ischemic heart disease, any heart failure, any sequelae or late effects of cerebrovascular disease, hypothyroidism, prescription or diagnosis for hypertension, weight loss or muscular wasting/atrophy, renal failure, any possible non-bipolar antiepileptic mood stabilizer indication recorded in week prior to/at cohort entry date, bipolar disorder subtype (Type I or Type II/not specified), bipolar disorder severity (crisis, active, psychosis or acute care, active outpatient, non-active or in remission), number of other psychiatric conditions (factor: 0, 1, or ≥2 of other depression excluding bipolar-type, anxiety disorders, PTSD, schizophrenia spectrum disorder), depressed episode as most recent bipolar disorder episode, schizophrenia or schizoaffective disorder, mania, number of primary position bipolar disorder diagnoses, number of primary diagnoses for any psychiatric-related condition (square root transformation), number of days in psychiatric hospitalization or partial hospitalization (square root transformation), any active bipolar disorder (mania, depression, psychosis) recorded at cohort entry, inpatient or emergency department admission for psychosis or related during baseline, any inpatient care for psychiatric or substance use disorders, hospitalization for bipolar disorder (1st or 2nd diagnosis position), most severe neurocognitive impairment recorded at baseline (suspected prodromal MND, MCI, mild-moderate ADRD), number of ADRD diagnoses, primary position ADRD diagnosis, unspecified amnestic disorder or memory loss, dementia in other diseases classified elsewhere, any possible ADRD complication or treatment (symbolic dysfunction, agitation or aggression, other behavioral disturbance, abnormal gait or lack of coordination, dysphagia or prescription for donepezil, galantamine, rivastigmine, or memantine), diagnosis for MND in week before/at cohort entry, use of any antidepressant excluding medications for bipolar depression, use of opioids or related analgesics, use of non-mood stabilizer anticonvulsants, use of statins or other lipid-lowering medications, use of oral antidiabetics, use of antiplatelets or non-steroidal anti-inflammatory drugs, number of distinct prescriptions during baseline, number of cardiovascular medication classes during baseline (antihypertensives, statins, antiplatelets, nitrates, oral anticoagulants), highest-intensity utilization recorded at cohort entry (acute inpatient or emergency department, non-acute setting, other outpatient, prescription only), index prescription from specialty or facility pharmacy, number of days with home health care (modeled with a natural logarithm transformation), home health visit in week before/at cohort entry, home health visit not preceded by hospitalization in prior 30 days, any claim for durable medical equipment during baseline.

We adjusted for more parsimonious sets of potential confounders in the observational emulations with MarketScan and Optum Clinformatics Data Mart databases, and within subgroup and sensitivity analyses (**Suppl. S6**). All potential confounders were well-balanced between treatment groups after weighting by IPTWs, with all absolute standardized differences <0.1 for the main Medicare analysis and all absolute standardized differences <0.2 for other analyses. Extended tables of baseline characteristics are provided in **Suppl. S11**.

#### Sensitivity analyses

To evaluate the robustness of our results, we conducted nine prespecified sensitivity analyses (SAs), comprising modifications of eligibility criteria and outcome definitions. First, we excluded patients with any parkinsonisms, hip fracture, or post-stroke paresis during baseline, which were considered potential causes of complicated dementia unrelated to ADRD progression (SA1). Second, we excluded patients with a diagnosis for any other possible indication for an antiepileptic mood stabilizer in the week before or on the day of treatment start, including epilepsy or convulsions, other parkinsonisms, migraine, trigeminal neuralgia, neuropathies and related nerve pain, schizophrenia, agitation, or multiple sclerosis (SA2). Third, we used a more stringent definition for bipolar disorder, requiring at least two diagnoses with specified ICD codes on two separate days in the 12-month baseline period (SA3). Fourth, we used a more stringent definition for MND at baseline, requiring two diagnoses with specified ICD codes on two separate days in the 12 months baseline period (SA4). Fifth, we evaluated composite outcomes for ADRD progression or death to account for death as a potential competing event (SA5). Sixth, we shortened the treatment washout period to 6 months, which may better represent real-world treatment patterns for patients with bipolar disorder (SA6). We also used several alternate specifications of the main outcome definition to address possible misclassification, including restricting to a diagnosis in the primary position (SA7), requiring a primary or admitting diagnosis in inpatient or long-term care settings (SA8); and using an alternative algorithm^17^ requiring a diagnosis for progression to advanced ADRD followed by an incident prescription (180 day washout) for medications used to treat ADRD symptoms (donepezil, galantamine, rivastigmine, or memantine) within the next 90 days (SA9).

In post-hoc sensitivity analyses, we removed three less specific ICD-9 diagnosis codes for dementia with delirium from the main outcome definition^17^ (SA10) and defined several alternate specifications of outcomes that incorporated long-term care components (SA11), including: a) long-term care stay (at least 100 days) without ADRD requirement; b) long-term care stay (at least 100 days) with diagnosis for advanced (severe or complicated) ADRD; c) any suspected long-term care admission with ADRD without any duration requirement; and, d) any suspected long-term care admission with advanced ADRD without any duration requirement.

#### Validations in information-rich subsets of Medicare beneficiaries

Ascertainment of neurocognitive status is known to be challenging in claims databases. Using linked assessment data available for an information-rich subset of Medicare beneficiaries, we conducted three validations to assess the reliability of claims-based criteria used for eligibility, confounding adjustment, and outcome ascertainment.

Validations leveraged detailed cognitive and functional assessments from three supplementary sources: the Patient Assessment Instrument administered in inpatient rehabilitation facilities (IRF-PAI), the Outcome and Assessment Information Set (OASIS) administered by home health agencies, and the Minimum Data Set administered in nursing facilities. Assessments from the IRF-PAI and OASIS were used for evaluating the adequacy of claims-based eligibility criteria and confounding adjustment at baseline, prior to treatment initiation. (Because we excluded patients with pre-treatment nursing facility utilization, Minimum Data Set assessments were not available for included patients at baseline.) The Minimum Data Set was used to evaluate the claims-based criteria for ADRD progression during follow-up.

#### Validation Q1: Do claims-based eligibility criteria adequately identify mild neurocognitive disorder while excluding advanced ADRD?

To assess if claims-based eligibility criteria adequately identified MND while excluding advanced ADRD, we examined how many patients in the Medicare emulation met criteria for severe dementia at baseline based on IRF-PAI and OASIS instruments. If the diagnosis codes used to identify patients with MND and to exclude those with advanced ADRD at baseline do not reliably capture neurocognitive severity, we may inadvertently include patients who had already progressed to advanced ADRD. These cases would represent unrecorded, latent outcomes at baseline.

For patients with more than one PAI or OASIS assessment during the baseline period (the 12 months preceding treatment initiation), the most recent assessment was used. OASIS records were restricted to home health assessments administered at start of care, resumption of care, or discharge from care, and we excluded assessments occurring within 14 days after acute hospitalizations (via OASIS item M1000).

#### Severe dementia algorithm

Following the algorithm used by Anand et al,^21^ we defined severe dementia as severe cognitive impairment (IRF-PAI modified Cognitive Functioning Scale^27^ >=3 or OASIS Cognitive Function item M1700/M0560 ≥3), dependencies in three basic activities of daily living (toileting, bathing, dressing), and at least one dependency in an advanced activity of daily living (incontinence, speech, feeding/eating, transferring, walking/ambulation). Algorithm details are provided in **Suppl. S5.** We separately reported results from the IRF-PAI modified Cognitive Function Scale, IRF-PAI Brief Interview for Mental Status^26,28^ and OASIS Cognitive Function item M1700/M0560 as less specific measures of advanced neurocognitive impairment (**Table S13**).

#### Additional dementia severity measures

We also developed algorithms that combined available cognitive and functional impairment items from OASIS and IRF-PAI to obtain other proxy measures of dementia severity.^39^ Moderately severe to severe dementia was defined as an IRF-PAI modified Cognitive Function Scale^27^ or OASIS Cognitive Functioning item ≥3, impairment in three basic activities of daily living (showering, bathing, toileting) and dependence in at least one advanced activity of daily living (incontinence, speech, feeding/eating, transferring, walking/ambulation). Moderate *or* severe dementia was defined as at least moderate cognitive impairment (IRF-PAI modified Cognitive Function Scale ≥3, IRF-PAI Brief Interview for Mental Status of 0-12, or OASIS Cognitive Functioning item ≥3) plus impairments in three basic activities of daily living. Any (suspected) dementia was defined as the presence of both cognitive and functional impairments, operationalized as any cognitive impairment from OASIS or IRF-PAI items and at least one impairment in any activity of daily living, including assistance or dependence in basic, advanced, or instrumental activities (see **Suppl. S5**).

#### Validation Q2: Is there residual confounding in baseline cognitive status across treatment groups?

To assess the potential for residual confounding in cognitive status at baseline, we examined whether the distribution of cognitive function instruments differed between lithium and antiepileptic mood stabilizer initiators after adjustment with IPTWs. Validation Q2 used the same baseline assessment data and information-rich subset as Validation Q1. To evaluate whether claims-based confounding adjustment was satisfactory, we assessed covariate balance for different IRF-PAI and OASIS items after applying IPTWs computed from claims-based factors only.

We combined separate cognitive function instruments from IRF-PAI and OASIS to obtain one measure of baseline cognitive impairment (Figure 4C), while also computing balance for individual instrument components (**Table S14**). In addition to the cognitive and functional status measures from validation Q1, we extracted and evaluated balance between treatment groups for additional OASIS questionnaire items, including: documented memory deficit, documented impaired decision making, speech and oral expression performance, and ability to understand verbal content.

Absolute standardized differences were computed using pooled standard deviations and proportions from the overall population of lithium or antiepileptic mood stabilizer initiators, rather than the subset of patients with available OASIS/IRF-PAI data. This maintained a consistent denominator across different OASIS/IRF-PAI items and avoided introducing selection bias by conditioning on presence of a home health or inpatient rehabilitation facility encounter. However, it is important to note that validation items were derived from selected patient populations with available information from linked assessments and may not be representative of the overall patient population included in the emulation.

#### Validation Q3: Does claims-based ADRD progression during follow-up agree with impairment severity measured in nursing facility assessments?

Validation Q3 examined whether claims-based measures of ADRD progression correlated with validated instruments from the Minimum Data Set among patients admitted to nursing facilities during follow-up. We compared Minimum Data Set measures for cognitive and functional impairment with our two claims-based outcomes: progression to advanced (severe or complicated) ADRD and progression to long-term care stay with ADRD.

### Study population

The population for validation #3 comprised patients from the target trial emulation who were admitted to a nursing home or other nursing facility during the 5-year follow-up period (after treatment initiation and before disenrollment or death). We used the first available Minimum Data Set assessment during follow-up, which corresponded to an incident nursing facility admission given study exclusions for any nursing facility encounter during baseline.

### Minimum Data Set measures

Minimum Data Set measures were extracted from assessments administered within 30 days of the incident nursing facility admission. We implemented the Cognitive Function Scale^27^ as the primary measure of neurocognitive status. The Cognitive Function Scale is a hierarchical algorithm based on the Brief Interview for Mental Status (administered to patients who are able to actively participate) and the Cognitive Performance Scale (derived from staff assessments for patients unable to complete the Brief Interview for Mental Status). Both the Brief Interview for Mental Status and Cognitive Performance Scale are validated instruments with high sensitivity (83% and 94%, respectively) and specificity (92% and 94%) for cognitive impairment when compared with the modified Mini-Mental State examination.^26,28,29^ The Cognitive Function Scale combines these two instruments to obtain a consistent cognitive function measure for all nursing facility residents, ranging from 1 to 4 for (1) intact cognition, (2) mild impairment, (3) moderate impairment, and (4) severe cognitive impairment. While prior research^42,43^ has used a Cognitive Function Scale score ≥2 to indicate probable dementia, we assessed whether the Cognitive Function Scale correlated with our claims-based progression measures.

We also extracted results from the Brief Interview for Mental Status and Cognitive Performance Scale sub-components, and additional Minimum Data Set items for self-performance of daily living activities (**Suppl. S5**, **Table S14**). Because clinical staging for dementia considers how neurocognitive decline impacts daily functioning, we implemented additional algorithms proxying different levels of dementia severity based on co-occurring cognitive and functional impairments, guided by the Global Deterioration Scale.^39^ Moderately severe to severe dementia, moderate or severe dementia, any (suspected) dementia, and any cognitive impairment were identified using algorithms similar to validations Q1-Q2, mapped to Minimum Data Set data elements (see **Suppl. S5**).

### Validating claims-based measures for MND staging and the outcome of progression to advanced ADRD

The outcome of progression to advanced ADRD was defined as the transition from any baseline MND stage to advanced (severe or complicated) ADRD during follow-up. Validating this outcome therefore required evaluation of both the baseline staging of MND and the outcome ascertainment.

Patients in validation #3 were assigned a claims-based stage according to the most severe neurocognitive disorder diagnosis code recorded from the beginning of baseline (365 days prior to treatment initiation) through 30 days after the incident nursing facility admission.

Because patients were eligible to progress beyond MND during follow-up, this yielded four possible claims-based stages: suspected prodromal MND without MCI/ADRD < MCI without ADRD < mild-moderate ADRD without complications < advanced ADRD (see **Suppl. S4**). We assessed whether patients classified as having advanced ADRD in claims data showed more severe impairment on Minimum Data Set assessments relative to those without advanced ADRD. We also reported the distribution of Minimum Data Set items stratified by claims-based stage to determine whether claims-based diagnosis codes reliably proxied neurocognitive decline across the full continuum.

### Validating the outcome of long-term care stay with ADRD

To evaluate the claims-based outcome of a long-term care stay with ADRD, we assessed Minimum Data Set information among the subset of patients experiencing this outcome during follow-up. Of 697 patients with this outcome, 483 had corresponding Minimum Data Set assessments at their first outcome-qualifying long-term care admission. Minimum Data Set criteria were assessed through 30 days after the later of either the 100th day in the nursing facility stay or the overlapping ADRD diagnosis. Characteristics for this population are shown in **Table S15** and Figure 5.

### Ethics and Reporting

The study was preregistered (Open Science Framework, https://osf.io/ug5ca/?view_only=39fa39343a4e49e698044014613d28e7), and minor adjustments to the initially preregistered protocol are reported in **Table S2**. This study was approved by the Brigham and Women’s Hospital institutional review board and follows reporting guidelines for Transparent Reporting of Observational Studies Emulating a Target Trial (TARGET)^44^ (**Table S3**).

### Software

Statistical analyses were performed using Aetion Substantiate (version 5) and R software (version 4.4.1).

## Data Availability

The authors are third-party users of US claims data (Medicare parts A, B, and D; Merative MarketScan; Optum Clinformatics Data Mart). The respective data use agreements and licensing agreements do not allow sharing of patient-level data with third parties. However, interested parties can contact the authors and request replication analyses.

## Acknowledgments

We thank Ryan Dimentberg, MD, MBE, for helpful insights on clinical management of bipolar disorder and neurocognitive disorders.

## Funding

N.K. received support from the German Heart Foundation (S/02/24, SRF-HF/24, RWE/12/25). A.R.W. received training support from the National Institutes of Health (T32-HL098048). S.S. receives funding from the National Institutes of Health (R01-HL141505, R01-AR080194). R.J.D. reports serving as Principal Investigator on a research contract from National Institutes of Health related to ADRD (75N95019C00057).

## Author contributions

Concept and design: S.C and A.R.W. Acquisition, analysis or interpretation of data: all authors. Drafting of the manuscript: A.R.W. and S.C and R.J.D. Critical revision of the manuscript for important intellectual content: all authors. Statistical analysis: A.R.W. and S.C. Administrative, technical or material support: R.J.D. Supervision: R.J.D.

## Conflict of Interest Disclosures

A.R.W., S.C, G.D., P.W., and N.K have no conflicts of interest to disclose. S.S. reports personal fees from Aetion Inc, a software-enabled analytics company, and grants from Bayer, UCB, and Boehringer Ingelheim to Brigham and Women’s Hospital outside the submitted work. R.J.D. reports serving as Principal Investigator on research grants from Bayer and Vertex to the Brigham & Women’s Hospital for unrelated projects.

## Code availability

The analytic code (R scripts and Aetion workflows) used in this study is available from the corresponding author upon reasonable request.

---

**Extended Data Table 1.**
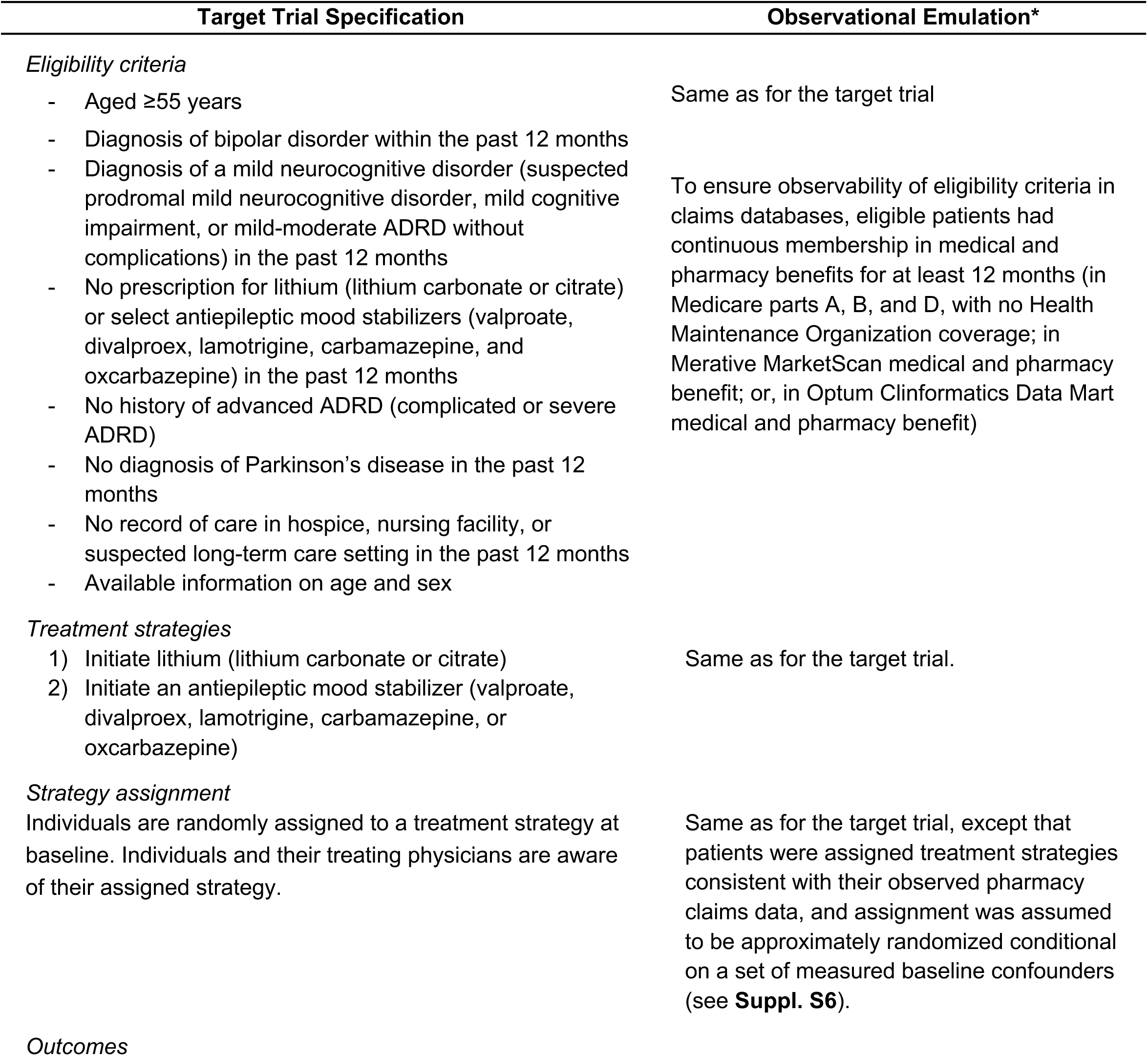
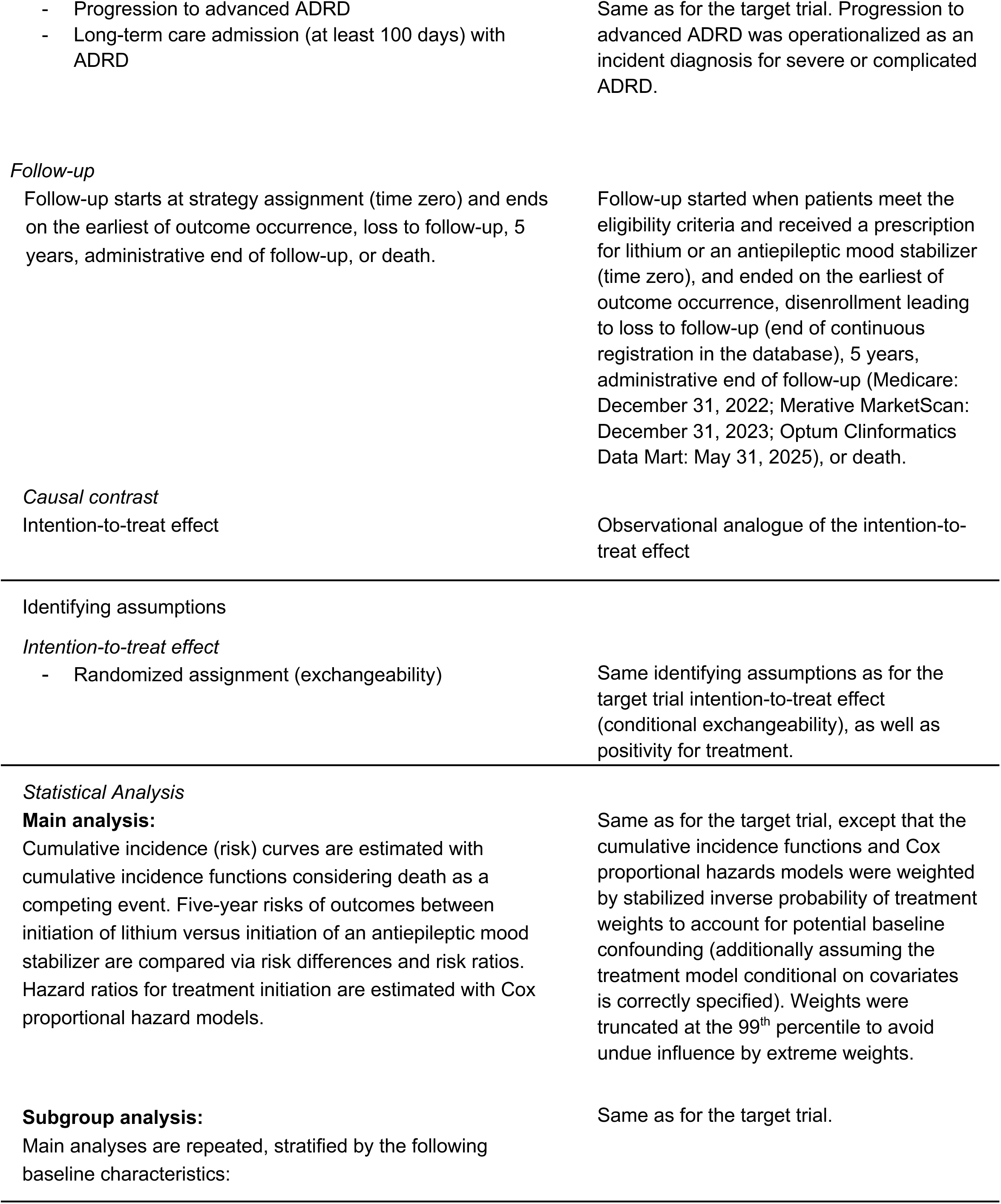
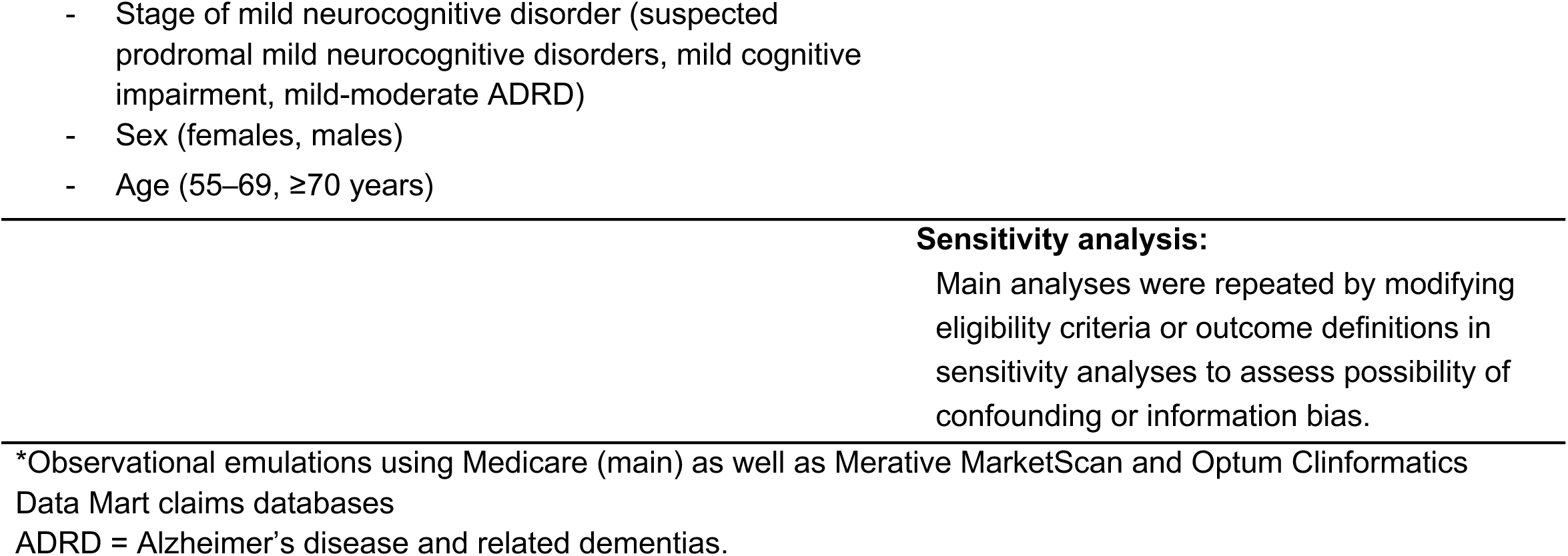
Specification of a target trial and observational emulation comparing initiation of lithium versus antiepileptic mood stabilizers in the progression of Alzheimer’s disease and related dementias among patients with bipolar disorder and a mild neurocognitive disorder

**Extended Data Figure 1.**
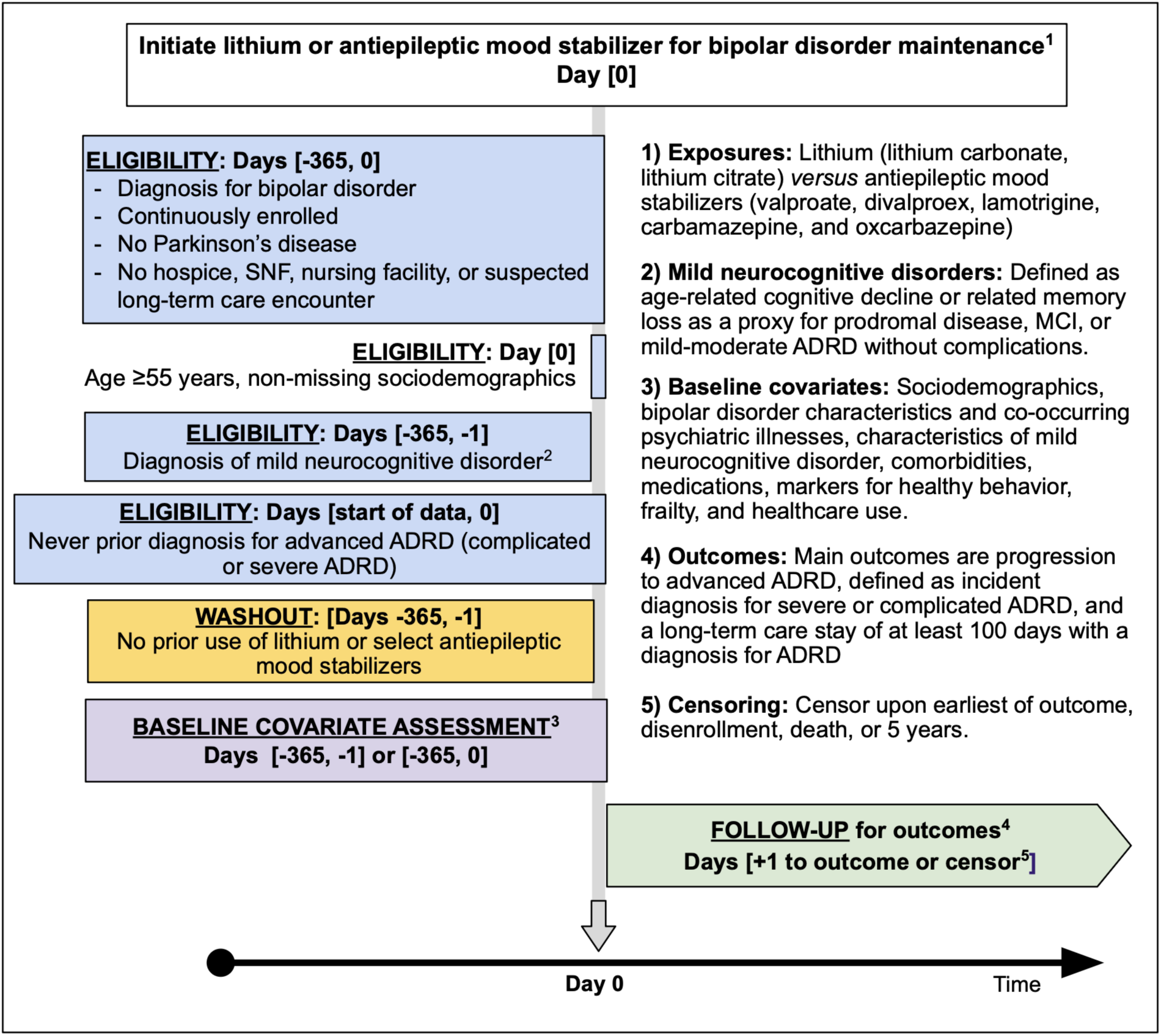
Study design diagram for observational emulations of target trial.

**Extended Data Table 2.** Key limitations, mitigation strategies, and supporting evidence.

| Potential threat | Rationale | Mitigation strategies | Supporting evidence |
| --- | --- | --- | --- |
| **Baseline misclassification** of neurocognitive disorder | If diagnosis codes used to include MND and exclude advanced ADRD are unreliable, this could result in the inclusion of individuals who have already progressed to advanced ADRD (unrecorded latent outcomes) or persons without any neurocognitive impairment. | • <u>Validation Q1</u>: Evaluated adequacy of eligibility criteria for identifying MND using linked data from home health and inpatient rehab encounters<br>• <u>Subgroup analyses</u> stratified by MND stage (suspected prodromal MND, MCI, mild-moderate ADRD)<br>• <u>SA1</u>: excluded other possible causes of cognitive issues (hip fracture, parkinsonism, post-stroke paresis)<br>• <u>SA4</u>: Required $\geq 2$ MND diagnoses | • <u>Validation Q1</u>: Both severe and moderately-severe dementia were very rare, and impairment indicators were mostly mild, supporting the reliability of diagnosis-based eligibility criteria.<br>• <u>Subgroup analyses</u>: Overall results are driven by risk reductions in MCI and ADRD subgroups, for which diagnosis codes may be more reliable (relative to the less specific suspected prodromal disease group).<br>• <u>SA1 and SA4</u>: Results consistent with main analysis |
| **Outcome misclassification** of ADRD progression | Outcome definition for ADRD progression relies on diagnosis sub-codes for severe ADRD or 'dementia with complication', which lack specific validation, and may not reliably indicate disease progression. Further, dementia complications during follow-up could be misattributed to bipolar disorder sequelae, or vice versa. | • <u>Validation Q3</u>: evaluated whether diagnosis-based outcome correlates with cognitive impairment among nursing facility admissions<br>• <u>Alternative outcome definitions</u> incorporated long-term care admissions as possibly more reliable proxies for disease progression, requiring prescription for ADRD symptoms (SA9), and excluding less specific dementia with delirium codes (SA10) | • <u>Validation Q3</u>: Diagnosis for advanced (severe or complicated) ADRD reliably proxies impairment severity, with severity correlating across levels (suspected prodromal MND < MCI < uncomplicated < advanced ADRD)<br>• <u>Alternative outcomes</u> are consistent with main findings, showing equal or larger lithium benefit relative to main outcome |
| **Residual confounding by indication** | Although antiepileptic mood stabilizers and lithium are both considered first-line bipolar disorder treatments, confounding by indication remains a possibility.<br>i) Clinical caution with lithium in severe kidney or heart disease may lead to preferential antiepileptic mood stabilizer use among sicker patients or as second-line therapy after lithium intolerance.<br>ii) Some antiepileptic mood stabilizer initiators may be treating non-bipolar disorder conditions (e.g., epilepsy, nerve disorders) or may be preferentially prescribed antiepileptic mood stabilizers to address both bipolar disorder and these conditions. Conversely, lithium may be used in patients with more severe bipolar | • <u>Principled new-user design with stringent exclusions</u>: required incident use and specific bipolar disorder diagnosis, exclude for any nursing facility care during baseline (off-label antiepileptic mood stabilizers for dementia is most common in nursing homes)<br>• <u>Robust adjustment strategy</u>: Assessed balance for >300 possible confounders (adjusted for >70), including bipolar disorder severity, psychosis history, co-occurring antiepileptic mood stabilizer indications, and cognitive status.<br>• <u>Validation Q1</u>: investigated possible misclassification of baseline cognitive status<br>• <u>Validation Q2</u>: evaluated residual confounding in baseline cognitive status<br>• <u>SA2</u>: excluded persons with diagnosis for non-bipolar disorder antiepileptic mood stabilizer indication in the week | • <u>Balance</u>: All covariates are well-balanced (ASDs<0.1) after adjustment, suggesting equipoise on observed covariates. Bipolar disorder and other possible antiepileptic indications are well-balanced across groups after adjustment (minimizing concern ii).<br>• <u>Validation Q1</u>: Provides reassurance that the cohort has adequately excluded persons with more advanced impairment/ADRD<br>• <u>Validation Q2</u>: Shows similar baseline impairment levels between antiepileptic mood stabilizer and lithium users<br>• <u>SA2 and SA3</u>: Results consistent with primary analysis |
|  | disorder.<br>iii) In a worst-case scenario, antiepileptic mood stabilizers may be prescribed off-label for dementia-related psychosis or agitation rather than bipolar disorder maintenance, though this would also require misclassification of baseline cognitive status (threat #1). | before/at cohort entry<br>• <u>SA3</u>: Require $\geq 2$ bipolar disorder diagnoses |  |
| **Footnote:** ADRD = Alzheimer's disease and related dementias, ASD = absolute standardized difference, BD = bipolar disorder, MCI = mild cognitive impairment, MND = mild neurocognitive disorder; SA = sensitivity analyses. |  |  |  |

## Notes

### Author Declarations

This study was approved by the Brigham and Women's Hospital institutional review board

